# The role of the spleen in red blood cell loss caused by malaria: a mathematical model

**DOI:** 10.1101/2025.05.08.25327032

**Authors:** Robert Moss, Saber Dini, Steven Kho, Bridget E. Barber, Pierre A. Buffet, Megha Rajasekhar, David J. Price, Nicholas M. Anstey, Julie A. Simpson

**Author notes:** These authors contributed equally to this work (Robert Moss, Saber Dini). These authors contributed equally to this work (Nicholas M. Anstey, Julie A. Simpson).

## Abstract

The human spleen significantly influences red blood cell (RBC) dynamics due to its ability to retain and/or remove RBCs from peripheral blood circulation. This filtering can mediate a range of malaria disease manifestations, depending on the physiological properties of the spleen. Data collected from patients undergoing splenectomy in Papua, Indonesia, revealed that in asymptomatic infections the spleen harboured substantially more infected RBCs than were circulating in the peripheral blood and that the spleen is also congested with uninfected RBCs. We hypothesise that two conditions hold for the spleen to retain such a high proportion of infected and uninfected RBCs: (i) the retention rate of RBCs is significantly higher than in uninfected patients; and (ii) phagocytosing macrophages cannot clear all of the infected RBCs from the spleen. In this paper, we present a mathematical model of RBC dynamics that includes, for the first time, the spleen as a compartment capable of retaining large numbers of infected and uninfected RBCs in *Plasmodium falciparum* and *P. vivax* infections. By calibrating the model to the Papuan data, we demonstrate that the spleen plays a significant role in removing not only infected RBCs but also uninfected RBCs. Uninfected RBC retention in the spleen, attributable to malaria, is substantially higher than circulating RBC loss due to parasitisation, for infections by both *Plasmodium* species. In chronic infections, the ratio of circulating uninfected RBCs lost to splenic retention per circulating uninfected RBC lost to parasitisation is 17:1 for *P. falciparum* and 82:1 for *P. vivax*. These ratios are larger than previously published estimates for acute clinical infections.

## 1 Introduction

The human spleen removes defective and senescent red blood cells (RBCs) from circulation. This removal is primarily facilitated by the narrow inter-endothelial slits through which RBCs must pass in order to return to the peripheral circulation (Groom et al., 1991). A RBC must be sufficiently deformable in order to cross these inter-endothelial slits and avoid biomechanical trapping and splenic clearance.

The spleen has long been considered a protective organ against *Plasmodium* infection, by removing dead/damaged intraerythrocytic parasites from the blood circulation after antimalarial treatment (Chotivanich et al., 2002). *Plasmodium falciparum* (Pf)-infected RBCs (iRBCs) have reduced deformability, and so are more likely to be retained in the spleen and subject to splenic clearance (Buffet et al., 2011). Moreover, in patients treated with artemisinins, “pitting” (a splenic process where parasite remnants are removed from iRBCs while crossing inter-endothelial slits) helps to recover large numbers of RBCs (Schnitzer et al., 1972). Consequently, studies have repeatedly shown that patients who have undergone a splenectomy (removal of the spleen) have higher risk of clinical malaria and are more prone to adverse outcomes (Israeli et al., 1987; Boone and Watters, 1995; Bach et al., 2005; Bachmann et al., 2009; Kho et al., 2018; Kambuaya et al., 2023).

Despite these protective benefits conferred by the spleen, Kho et al. (2021a) recently showed that the spleen can also act as a reservoir for large quantities of viable malaria parasites. The authors studied 15 asymptomatic patients who underwent splenectomy in Papua, and found that asexual parasitaemia in the spleen of these patients was, on average, 289-fold and 3590-fold higher than in peripheral circulation, for patients infected with Pf and *Plasmodium vivax* (Pv), respectively. This suggests a new paradigm: that the spleen can harbour a large parasite biomass and sustain chronic malaria infection. The study also revealed that in the majority of these asymptomatic individuals, the size of the spleens were enlarged (splenomegaly), and was later described to be mostly a result of congestion with uninfected RBCs (uRBCs) (Kho et al., 2023). Similarly, Woodford et al. (2021) observed an increase in splenic volume during the early stages of experimentally induced Pv infection. These studies and others highlight the importance of the spleen as a compartment for hidden malaria parasites, with major implications for our understanding of the biology, pathology, and treatment of malaria.

In endemic regions, asymptomatic and submicroscopic *Plasmodium* infections are associated with elevated risks of anaemia (Pava et al., 2016; Bahati et al., 2020) and congestion of the spleen has been identified as a major cause of apparent uRBC loss from circulation in asymptomatic infections (Kho et al., 2023). In acute clinical malaria, there is clear evidence that a large proportion of RBC loss is also attributable to uRBCs. Jakeman et al. (1999) fitted a mathematical model to historical data (albeit without inclusion of a splenic compartment) from neurosyphilis patients undergoing malaria therapy and inferred that parasitisation (invasion of uRBCs by merozoites) only caused around 10% of observed RBC loss. Analyses of human data from epidemiological studies and experimentally-induced blood-stage infections have also found that parasitisation accounts for only a small fraction of overall RBC loss (Price et al., 2001; Collins et al., 2003; Woolley et al., 2021).

Reduced uRBC deformability in patients with falciparum and vivax malaria has been reported (Dondorp et al., 1999; Rey et al., 2014; Ishioka et al., 2015; Barber et al., 2018; Rathnam et al., 2024), and increased splenic filtration stringency has also been proposed (Looareesuwan et al., 1987; Henry et al., 2022; Kho et al., 2023), both of which may contribute to increased splenic trapping of uRBCs and apparent loss in circulation. In a rat malaria model, Safeukui et al. (2015) found that increasing parasite removal in the spleen also increased uninfected-erythrocyte removal in the spleen. Furthermore, multiple studies in clinical malaria show that splenomegaly is associated with anaemia, including severe malarial anaemia (Giha et al., 2008; Cserti-Gazdewich et al., 2013; Kotlyar et al., 2014; Connon et al., 2021). Thus, like in asymptomatic infections, the spleen may play a major direct role in the development of anaemia in clinical disease.

In this study, we present a within-host mathematical model that characterises RBC dynamics in response to malaria, and includes the spleen as an explicit compartment that interacts with the peripheral circulation. The model accounts for RBC age-dependence in various regulatory processes that govern RBC dynamics. We use this model to quantify how splenic processes, such as the removal and retention of uRBCs and iRBCs from the circulation, and RBC phagocytosis by macrophages, may result in different manifestations of malaria. In particular, we identify parameter values for which the model reproduces the observed numbers of uRBCs and iRBCs in the spleen and circulation of asymptomatic Pf- and Pv-infected individuals with a large hidden splenic biomass (Kho et al., 2021a). By calibrating the model to these cross-sectional splenectomy data, we are able to characterise chronic asymptomatic infections, but stress that the model results do not accurately describe the acute infection phase (i.e., the transition from initial blood infection to chronic asymptomatic infection). We also use this model to quantify the ratio of circulating uRBCs lost to splenic retention per circulating uRBC lost to parasitisation, and report how our values differ from previous estimates without inclusion of a splenic compartment. Our findings highlight the spleen as a site in which major RBC retention and parasitisation can be achieved.

## 2 Materials and Methods

### 2.1 Patient data

Model outputs were compared to published uRBC and iRBC counts in the circulation and spleen of 15 asymptomatic adults in Papua, Indonesia (9 Pf infections, 6 Pv infections) who underwent splenectomy (Kho et al., 2021a). Splenectomy in this cohort was primarily due to splenic injury from trauma (8 Pf, 5 Pv); the remaining two patients (1 Pf, 1 Pv) underwent elective splenectomy due to clinically significant splenomegaly.

To define the RBC count at homeostasis, in the absence of malaria, we used published RBC counts (10^6^*/µl*) and blood volumes (L) from 708 healthy Papuans (negative results for both *Plasmodium* microscopy and PCR), who reported no fever within the preceding 24 hours, as collected in a cross-sectional household survey conducted in southern Papua, Indonesia (Pava et al., 2016).

### 2.2 Ethical approval

We obtained ethical approval from the Human Research Ethics Committee of Northern Territory Health and Menzies School of Health Research for (a) the uRBC and iRBC counts in 15 asymptomatic Papuan adults (HREC-2010-1397); and (b) the RBC counts and blood volumes from 708 health Papuans (HREC-2010-1434).

### 2.3 Mathematical model

The model structure is outlined in Figure 1, the model cell populations are described in Table 1, and rate parameters are described in Table 2. The model is implemented in the R programming language (R Core Team, 2021) as a set of difference equations that describe the evolution of the age-structured RBC populations over a series of one-hour time steps. We present here an overview of the processes that govern RBC dynamics in the model. For further details, see the supplementary Model Description (A).

**Figure 1:**
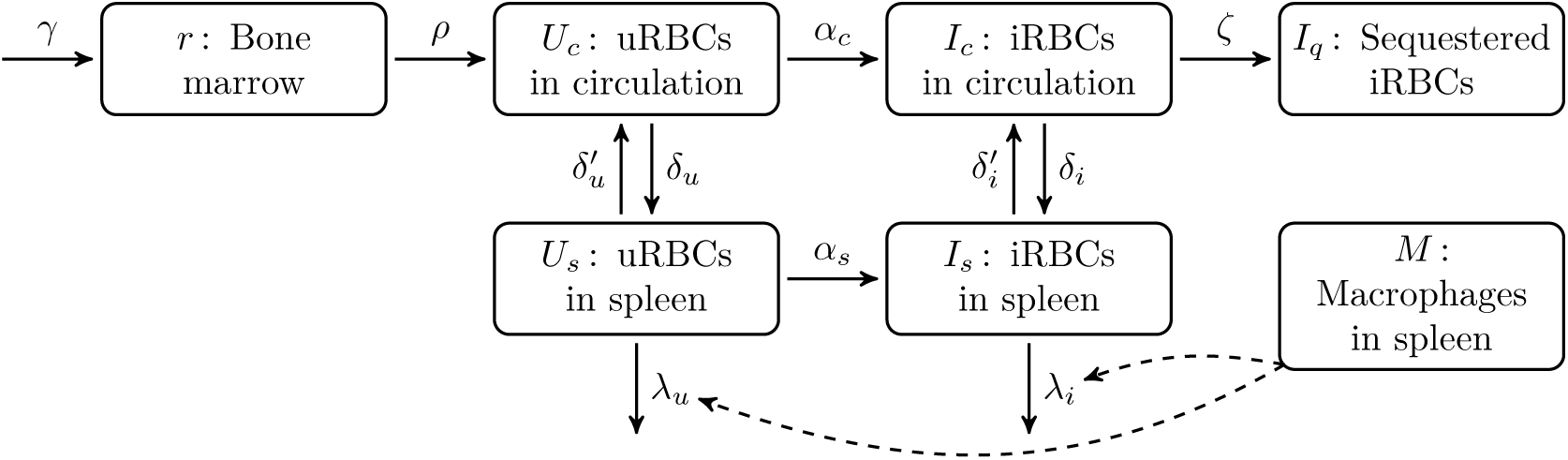
A schematic of the model structure, showing the various compartments in which red blood cells (RBCs) may reside, and the permitted movements of RBCs between these compartments. See Table 1 for descriptions of each cell population, and Table 2 for descriptions of each rate parameter. In brief, normoblasts are produced in the bone marrow and mature into reticulocytes, which are then released from the bone marrow into the circulation, from where they may pass through the spleen and either be retained or released back into the circulation. Uninfected RBCs (uRBCs) can be parasitised in the circulation and in the spleen. Infected RBCs (iRBCs) in the circulation can become sequestered in the microvasculature (Pf only). Phagocytosis of uRBCs and iRBCs in the spleen is driven by the splenic macrophage population (indicated by dashed lines).

**Table 1:** The cell populations in the model, with respect to age *a* and time *t* where appropriate.

| Symbol | Compartment | Cell type | Lifespan (days) |
| --- | --- | --- | --- |
| $r(a, t)$ | Bone marrow | Normoblasts and reticulocytes | 3.5 |
| $U_c(a, t)$ | Circulation | Uninfected RBCs | 120 |
| $I_c(a, t)$ | Circulation | Infected RBCs | 2 |
| $I_q(a, t)$ | Microvasculature | Infected RBCs | 2 |
| $U_s(a, t)$ | Spleen | Uninfected RBCs | 120 |
| $I_s(a, t)$ | Spleen | Infected RBCs | 2 |
| $M(t)$ | Spleen | Red-pulp macrophages | — |

**Table 2:** The rate parameters that govern the movements of RBCs between the model compartments.

| Symbol | Process |
| --- | --- |
| $\gamma$ | Production rate of normoblasts in the bone marrow. |
| $\rho$ | Release rate of reticulocytes from the bone marrow into the circulation. |
| $\delta_u$ | Removal rate of uRBCs from the circulation into the spleen. |
| $\delta'_u$ | Return rate of uRBCs from the spleen into the circulation. |
| $\lambda_u$ | Phagocytosis rate of uRBCs in the spleen. |
| $\lambda_u^M$ | uRBC phagocytosis rate per splenic macrophage. |
| $\delta_i$ | Removal rate of iRBCs from the circulation into the spleen. |
| $\delta'_i$ | Return rate of iRBCs from the spleen into the circulation. |
| $\lambda_i$ | Phagocytosis rate of iRBCs in the spleen. |
| $\lambda_i^M$ | iRBC phagocytosis rate per splenic macrophage. |
| $\alpha_c$ | Invasion rate of released merozoites in the circulation. |
| $\alpha_s$ | Invasion rate of released merozoites in the spleen. |
| $\zeta$ | Sequestration rate of iRBCs into the microvasculature. |

#### 2.3.1 RBC production and release into the circulation

Erythropoiesis was modelled by the hourly production of *γ* normoblasts in the bone marrow; other organs were assumed to make negligible contributions. We selected the baseline value of *γ* such that, in the absence of infection, the circulating RBC population remains constant. Note that the precise value of *γ* depends on the rate at which uRBCs are removed by the spleen. We assumed that erythropoiesis increases in response to decreases in the circulating RBC population (Hillman, 1969).

Under normal circumstances, erythroblasts in the bone marrow mature into normoblasts and then reticulocytes over a period of 3.5 days, before being released into the circulation and maturing into normocytes after 1 day in the peripheral blood (Hillman, 1969; Koepke and Koepke, 1986). With increasing anaemia, the maturation time in the bone marrow shortens to a minimum of 1 day, with a corresponding increase in the time taken to mature into normocytes in the peripheral blood (Hillman, 1969; Koepke and Koepke, 1986). In the model, reticulocytes are initially released very slowly into circulation (hourly release probability of 0.001). Upon reaching the release age, which depends on the circulating RBC population, reticulocytes are rapidly released into the circulation (hourly release probability > 0.9999).

#### 2.3.2 Uninfected RBC circulation and retention in the spleen

In the model, uRBC retention in the spleen arises from the combination of (a) the removal rate from circulation *δ_u_*; and (b) the return rate from the spleen 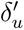. Note that any desired level of uRBC retention can be achieved with arbitrarily many combinations of removal and return rates, due to their inverse relationship (e.g., a high removal rate can be countered by a high return rate).

We primarily characterised uRBC retention in the spleen by the removal rate *δ_u_*. We assumed this rate is high for very mature rigid RBCs (110–120 days old) and for immature reticulocytes (*<* 4.5 days old), which have reduced deformability and increased cytoadherence capacity (Malleret et al., 2013) and will remain in the splenic red-pulp until they mature and are returned to the circulation (Kho et al., 2021b). RBCs that are 4.5–110 days old are removed from the circulation into the spleen at an extremely low rate. Accordingly, the uRBC return rate from the spleen into the circulation 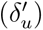 is zero for all uRBCs *except* maturing reticulocytes (3–4.5 days old).

Finally, we assumed that circulating iRBCs stimulate an increase in splenic retention of uRBCs, either from increased splenic filtration stringency (Looareesuwan et al., 1987; Henry et al., 2022; Kho et al., 2023) or reduced circulating uRBC deformability (Dondorp et al., 1999; Rey et al., 2014; Ishioka et al., 2015; Rathnam et al., 2024), with a maximum increase of 100% (i.e., a doubling of the removal rate *δ_u_*) when the circulating iRBC population *I_c_* exceeds 10^8^ cells.

#### 2.3.3 Infected RBC dynamics

Infected RBC retention in the spleen arises from the combination of (a) the removal rate from circulation *δ_i_*; and (b) the return rate from the spleen 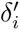. As per uRBC retention, there is an inverse relationship between these two rates. Safeukui et al. (2008) conducted *in vitro* experiments that showed 11% of Pf rings and 20% of Pf schizonts are retained in the spleen in every passage of the iRBCs through the spleen. Accordingly, we assumed that the removal rate increases with maturity (hourly removal probabilities of 0.43 for rings and 0.67 for schizonts), while the return rate decreases with maturity (hourly return probabilities of 0.017 for rings and 0.011 for schizonts).

We also assumed that circulating iRBCs stimulate an increase in splenic retention of iRBCs, with a maximum increase of 300% (i.e., a four-fold increase) when the circulating iRBC population *I_c_*exceeds 10^11^ cells.

For Pf infections, circulating iRBCs are also sequestered into the microvasculature at an age-dependent rate *ζ*. This rate is very low for Pf rings, with an hourly sequestration probability of ≪ 0.01, and increases with maturity up to a maximum hourly sequestration probability of 0.99 for schizonts.

#### 2.3.4 Infection of RBCs

Merozoites are released when iRBCs rupture 48 hours after parasitisation, and we assume that each ruptured iRBC infects 8 uRBCs (a parasite multiplication factor of 8). Merozoites released into the circulation infect uRBCs in the circulation, while we assume that merozoites released into the spleen *primarily* infect uRBCs in the spleen, but allow a small proportion of these merozoites (those from rupturing schizonts in the splenic perifollicular zones, or those escaping through inter-endothelial slits (Kho et al., 2021b)) to infect uRBCs in the circulation.

#### 2.3.5 RBC destruction in the spleen

Each individual macrophage destroys uRBCs at a rate 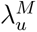 and destroys iRBCs at a rate 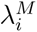 , and we assume that 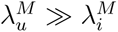 from the larger number of aged uRBCs compared to iRBCs that are retained and subject to phagocytosis at interendothelial slits in the spleen. The net phagocytosis rates *λ_u_* and *λ_i_* depend on the splenic macrophage population in the red-pulp *M* , which at steady-state is calculated from published healthy controls (Urban et al., 2005), and which expands during infection in proportion to the quantity of RBCs retained in the spleen (based on a higher red-pulp macrophage count in asymptomatic infections (Kho et al., 2023)). We assume there is no phagocytosis of uRBCs or iRBCs in the peripheral circulation.

#### 2.3.6 Differences between Pf and Pv

Pf can be sequestered into the microvasculature (*ζ >* 0) while Pv cannot (*ζ* = 0). The only other difference between the two species in this model is the age-dependent merozoite preference *β*(*a*) for uRBCs of age *a*, as illustrated in Figure S10 (supplementary materials). Pv parasites only invade reticulocytes and prefer immature reticulocytes (Malleret et al., 2015), whereas Pf invades RBCs of all ages, with some preference for younger RBCs (Wilson et al., 1977; Simpson et al., 1999).

Values for other biological parameters in the model are likely to differ between Pf and Pv, such as the parasite multiplication factor and the rate of iRBC removal from circulation due to differences in iRBC deformability. However, for simplicity we assumed identical values for these parameters for the two species. As the results of the sensitivity analysis show, the model dynamics are not particularly sensitive to the assigned values.

#### 2.3.7 Fitting and sensitivity analysis

The model comprises 34 parameters. Baseline values are listed in the supplementary Model Description (A). We were unable to estimate the model parameters using standard model-fitting approaches, because longitudinal circulation and splenic RBC data were not available. Instead, previously-published studies were used to derive many parameter values, and key parameters were systematically varied within biologically plausible ranges to identify values that could explain the reported cross-sectional splenectomy data, under the assumption that these data characterised the steady-state dynamics of chronic asymptomatic malaria infections. Further, since we used subject-level data from two different cohorts (see subsection 2.1), we calibrated the model against whole cohorts rather than individual subject data.

To quantify how well the steady-state model iRBC populations *I_c_* and *I_s_* matched the iRBC counts 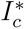 and 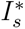 in the cross-sectional splenectomy data, we scaled the absolute error between the model and median datum by the spread of the splenectomy data:

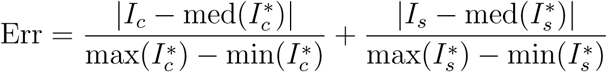

In the sensitivity analysis we varied all 34 model parameters by ±20%, using Latin hypercube sampling to draw 1000 samples for each parameter, and ran model simulations for Pf and Pv infections. We reported median-centred 50% and 95% uncertainty intervals, which correspond to the 0.25–0.75 and 0.025–0.975 quantile intervals, respectively.

## 3 Results

We begin by identifying plausible parameter combinations for which the model produces substantial splenic retention of iRBCs. We then demonstrate that, for some of these parameter combinations, the model is capable of producing chronic malaria infections that are consistent with the cross-sectional splenectomy dataset. For these parameter combinations, we show that uRBC retention in the spleen (attributable to malaria) is substantially higher than parasitisation of circulating uRBCs, for both Pf and Pv infections. Finally, we demonstrate that the model dynamics are not particularly sensitive to the choice of model parameter values, and that our primary findings are robust to the limits of our model calibration.

### 3.1 Retention of infected RBCs in the spleen

A key characteristic in chronic *Plasmodium* infections is that the large majority of iRBCs are found in the spleen and a minority are present in the circulation (Kho et al., 2021a). Here, we have identified the model transitions that have the greatest influence on iRBC retention in the spleen:

1. The removal rate of iRBCs from the circulation *δ_i_*, governed by the model parameter *δ_iR_* (removal rate of ring iRBCs); and
2. The phagocytosis rate of iRBCs in the spleen *λ_i_*, governed by the model parameter 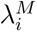 (iRBC phagocytosis rate per individual macrophage).

We systematically varied these two parameters, and observed that removal rates *δ_iR_* ≥ 0.1 result in chronic infections where the majority of iRBCs are retained in the spleen (Figure 2). The absolute removal rates of uRBCs and iRBCs into the spleen increase with the circulating parasite load, from baseline values for ≤ 10^7^ circulating iRBCs, up to a four-fold increase for ≥ 10^11^ circulating iRBCs. The removal rate *δ_iR_* = 0.1 corresponds to hourly removal probabilities of 0.095 (at baseline) to 0.33 (high parasite load).

**Figure 2:**
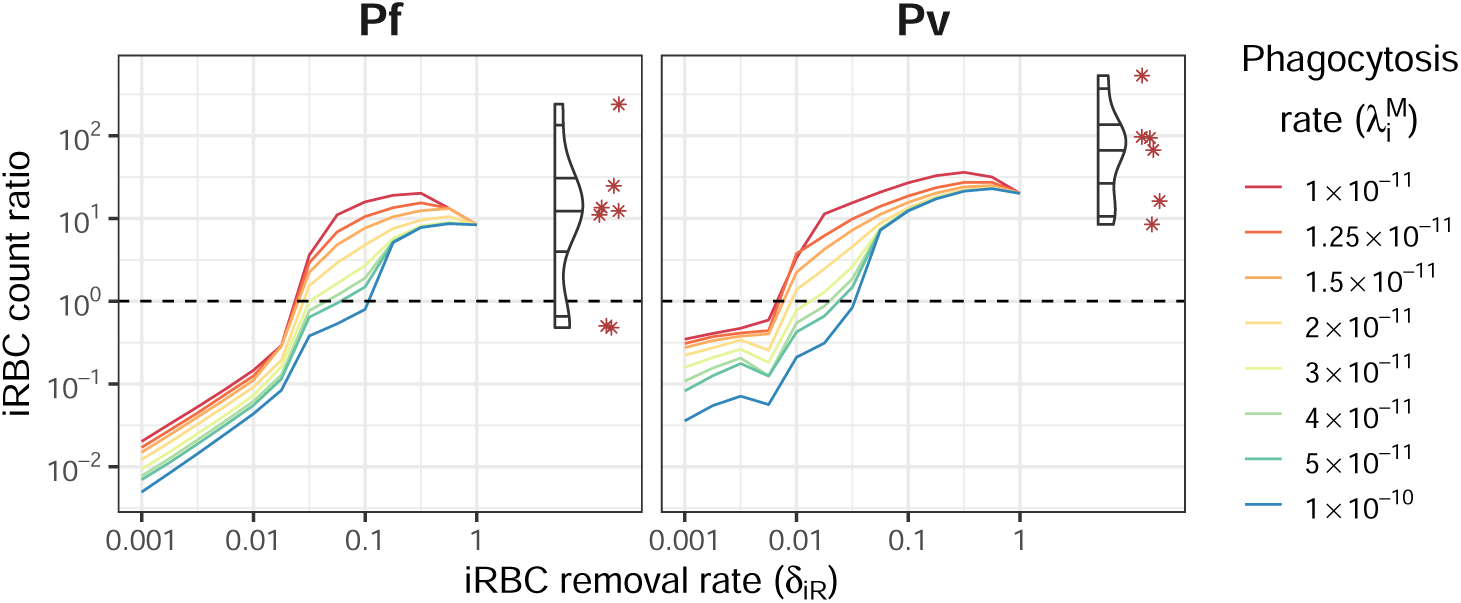
The steady-state ratio of iRBCs retained in the spleen (absolute counts, *I_s_*) to iRBCs in the circulation (absolute counts, *I_c_*), for a range of values for iRBC removal from circulation (*δ_iR_*) and iRBC phagocytosis in the spleen 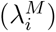, shown as coloured lines. Corresponding data in the splenectomy patients (Pf: n=9; Pv: n=6) are shown on the right of each plot for comparison (violin plots for distribution and red asterisks for observed data points).

### 3.2 Comparison to splenectomy patient data

A comparison of the model results to the absolute counts of peripheral and splenic uRBC and iRBC counts observed in the splenectomy patients is shown in Figure 3. While the circulating and splenic uRBC counts were relatively stable for removal rates *δ_iR_* ≥ 0.05 across a wide range of phagocytosis rates, the iRBC counts exhibited much greater sensitivity to both parameters.

**Figure 3:**
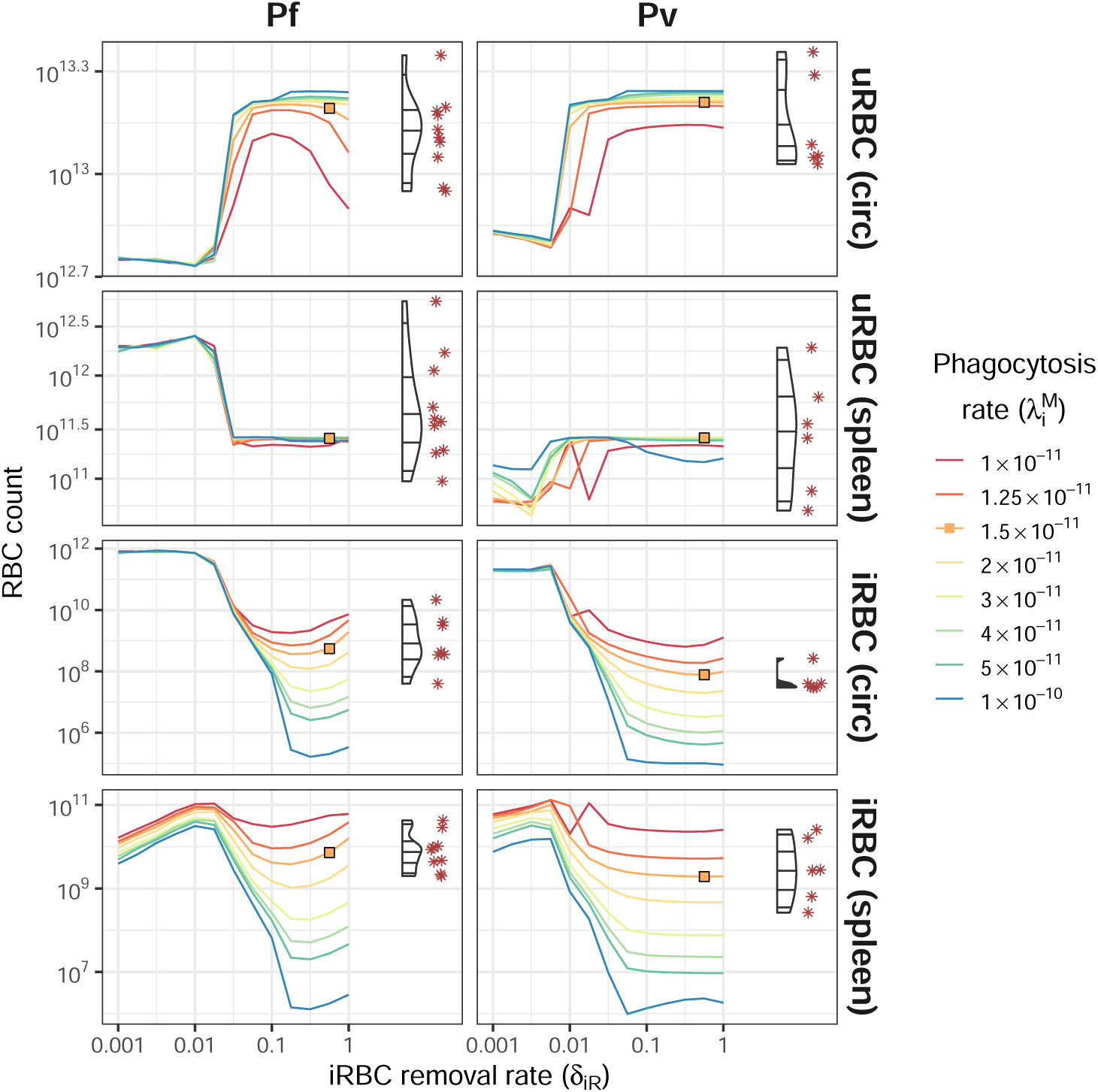
Steady-state uRBC and iRBC absolute counts in the circulation and spleen, for a range of values for iRBC removal from circulation (*δ_iR_*) and iRBC phagocytosis in the spleen 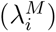, shown as coloured lines. Corresponding data in the splenectomised patients (Pf: n=9; Pv: n=6) are shown on the right of each plot for comparison (violin plots for distribution and red asterisks for observed data points). Orange squares indicate good agreement between the model and the data.

Based on the iRBC count ratio results (Figure 2), we considered parameter combinations where *δ_iR_* ∈ [0.1, 1]. For Pf infections, the best agreement with the clinical data was obtained with 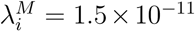 and *δ_iR_* ∈ [0.3, 0.6]. For Pv infections, the best agreement was obtained with 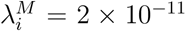 and *δ_iR_* ∈ [0.1, 0.6]. In Figures 3–5 we highlight one parameter combination that resulted in good agreement with the data for both species (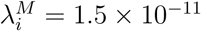, *δ_iR_*= 0.562).

In 11 individuals with asymptomatic infections, less than 5% of total-body RBCs were retained in the spleen (Pf: 6 of 9; Pv: 5 of 6) (Kho et al., 2021a). The model yielded similar steady-state proportions for iRBC removal rates *δ_iR_* ≥ 0.1, as shown in Figure 4.

**Figure 4:**
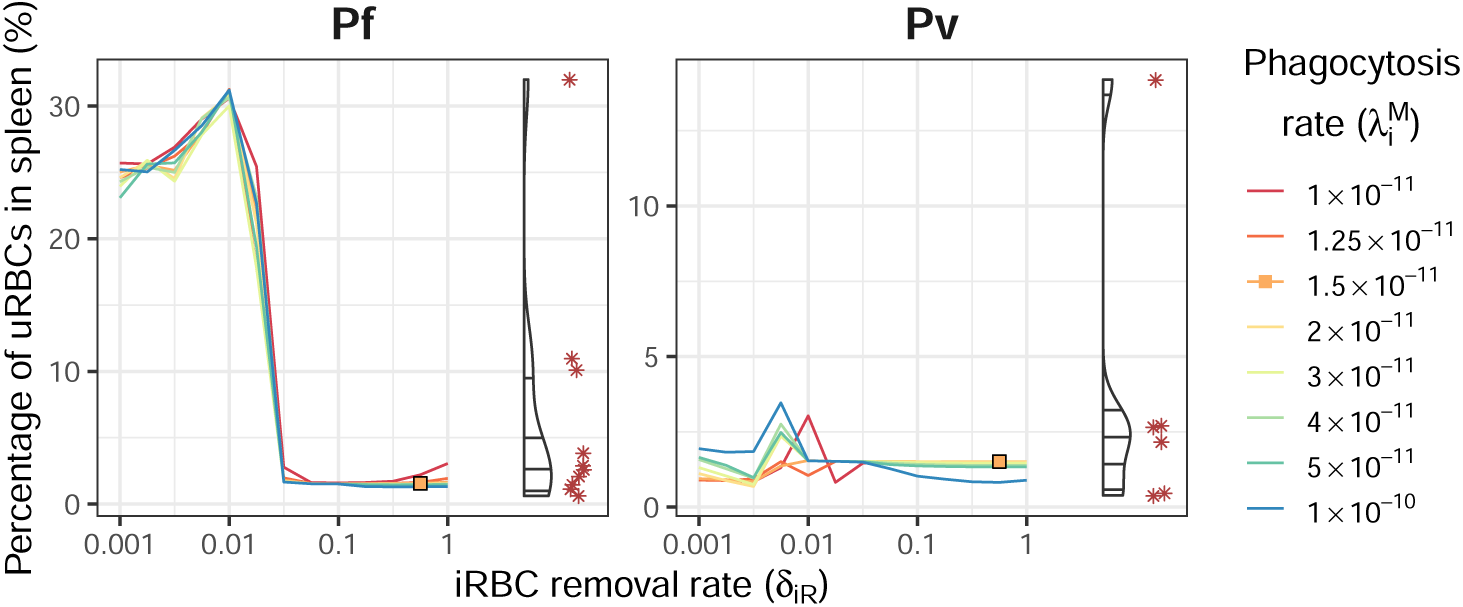
The steady-state percentage of total-body uRBCs that are retained in the spleen (*U_s_*), for a range of values for iRBC removal from circulation (*δ_iR_*) and iRBC phagocytosis in the spleen 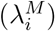, shown as coloured lines. Corresponding data in the splenectomised patients (Pf: n=9; Pv: n=6) are shown on the right of each plot for comparison (violin plots for distribution and red asterisks for observed data points). Orange squares indicate good agreement between the model and the data.

The remaining 4 individuals had massively enlarged spleens containing 10–32% of total-body RBCs, with the model yielding similar steady-state proportions for Pf infections at substantially lower iRBC removal rates *δ_iR_* ≤ 0.03, also shown in Figure 4.

These lower iRBC removal rates result in extreme anaemia, with haemoglobin levels as low as 5 g/dL (shown in Figure 5), and extremely high parasite levels (Figure 3). Lower iRBC removal rates also caused the model dynamics to become particularly sensitive to the rates of uRBC and iRBC retention, and phagocytosis, as evident in Figure 4 for *δ_iR_* ≤ 0.03.

**Figure 5:**
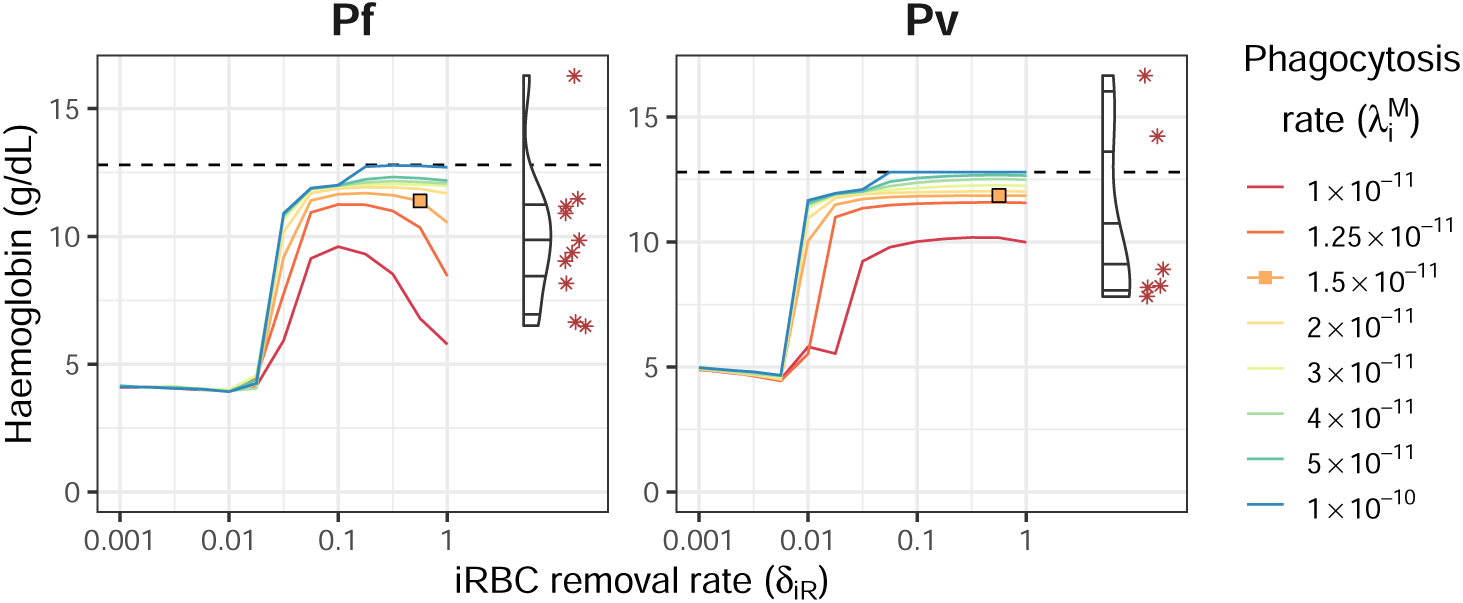
Steady-state haemoglobin, for a range of values for iRBC removal from circulation (*δ_iR_*) and iRBC phagocytosis in the spleen 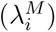, shown as coloured lines. Corresponding data in the splenectomised patients (Pf: n=9; Pv: n=6) are shown on the right of each plot for comparison (violin plots for distribution and red asterisks for observed data points). Orange squares indicate good agreement between the model and the data. Dashed lines indicate the baseline haemoglobin level of 12.8 g/dL for endemic healthy controls (Pava et al., 2016).

### 3.3 Initial and chronic infection stages

There are two processes in the model that remove uRBCs from the circulation: parasitisation by merozoites released by mature iRBCs, and retention of uRBCs in the spleen. In the model, the presence of iRBCs in the circulation results in a fold increase in uRBC removal from the circulation, and we can calculate the increase in uRBC retention in the spleen that is caused by infection.

As shown in Figure 6 (left panel), this malaria-associated retention of uRBCs in the spleen is substantially greater than the direct loss of circulating uRBCs due to parasitisation. In the early progressive stages of the model simulations, parasitisation causes only 0.01% of uRBC loss for Pf and only 0.02% of uRBC loss for Pv. In chronic infections (day 20 onwards), parasitisation causes 5.5% of uRBC loss for Pf and 1.2% of uRBC loss for Pv, and the ratio of circulating uRBCs lost to splenic retention per circulating uRBC lost to parasitisation is 17:1 for Pf and 82:1 for Pv.

**Figure 6:**
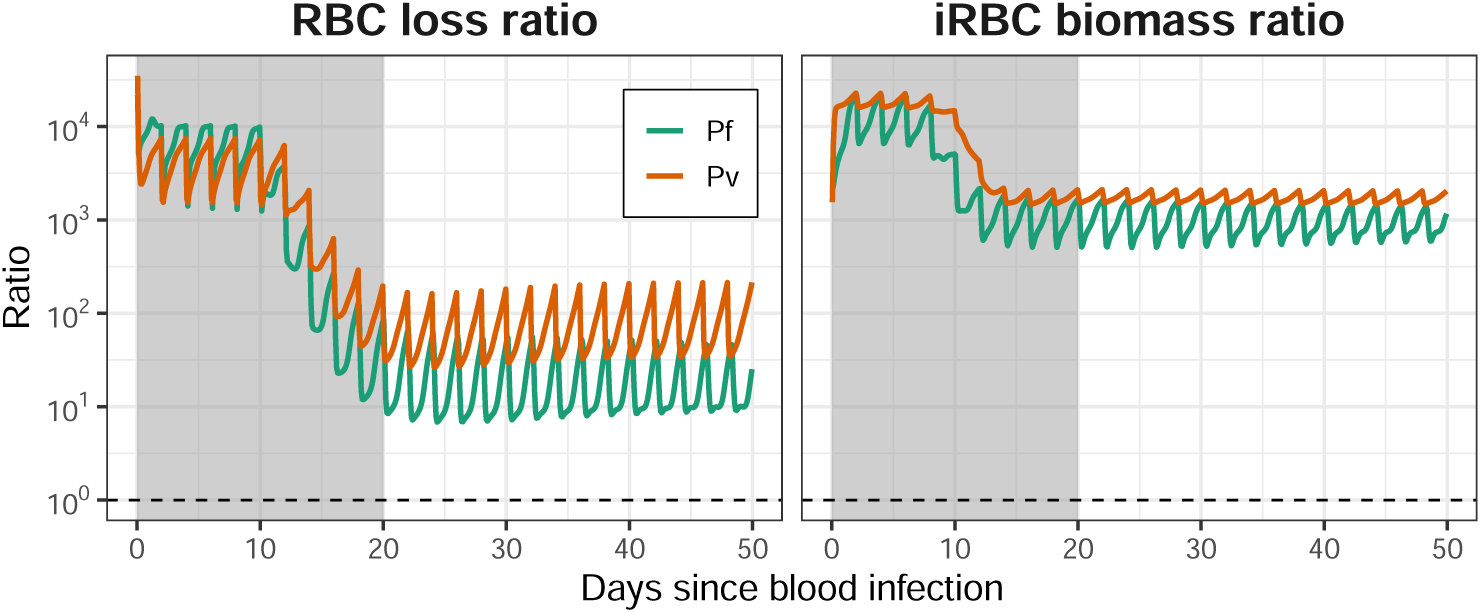
Left panel: the ratio of circulating uRBC loss due to increased retention in the spleen, versus the loss from circulation due to infection by malaria parasites. Right panel: ratio of iRBC biomass in the spleen to the iRBC biomass in the circulation. Dashed lines indicate ratios of 1:1. Shaded intervals indicate the acute infection phase (i.e., the transition from initial infection to chronic infection), which the model dynamics do not capture.

The ratio of iRBC biomass (the percentage of RBCs that are parasitised) in the spleen versus in the circulation is shown in Figure 6 (right panel). This ratio has been used by, e.g., Kho et al. (2021a) to characterise splenic tropism. The chronic steady-state ratios are 918:1 for Pf and 1701:1 for Pv; these values are similar orders of magnitude to those reported by Kho et al. (2021a) (Pf: mean 289, range 18–1530; Pv: mean 3590, range 2300–4210).

Recall that uRBC and iRBC phagocytosis rates in the model are directly proportional to the splenic macrophage population, and that increased RBC retention in the spleen stimulates an increase in the macrophage population. Relative to homeostasis, the phagocytosis rates are 11 times higher in chronic Pf and Pv infections (day 20 onwards).

### 3.4 Sensitivity analysis

We varied all 34 models parameters by ±20%. As shown in Figure A3, the model dynamics are not particularly sensitive to our reference parameter values. This is a positive feature of the model: many of these parameters cannot be estimated from (or indirectly informed by) experimental or clinical data, and this provides reassurance that the model dynamics are not substantially influenced by moderate adjustments to our reference values.

Across the 1000 model simulations for each species, the infection dynamics were extremely similar and the chronic steady-states remained consistent with the patient data. Accordingly, the iRBC biomass ratios (Figure A4) and uRBC loss ratios (Figure A5) also exhibited only modest variation across all of the model simulations.

## 4 Discussion

To our knowledge, this is the first mathematical model of Pf and Pv infections that describes the dynamics of uninfected and infected RBCs with inclusion of the spleen as a separate compartment where RBCs can be retained and phagocytosed. In this model, the spleen significantly influences whole-body RBC dynamics and the relationship between parasitaemia (parasite counts in the circulation) and non-circulating parasite load. Through simulations using biologically plausible parameter values, we were able to produce chronic malaria infections where the spleen retained an overwhelming majority of iRBCs, and also retained large quantities of uRBCs. This vision is consistent with early speculations triggered by the observation of innate retention of ring-iRBC in human spleens *ex vivo* (Safeukui et al., 2008), pointing to stringent retention in chronic Pf infections (Buffet et al., 2009). Our results are also consistent with experimental observations in the spleen and peripheral blood of asymptomatic adults naturally infected with Pf and Pv in Papua, Indonesia (Kho et al., 2021a, 2023).

Our model simulations showed that large quantities of parasites can be retained in the spleen when the iRBC retention rate is sufficiently high, and the splenic phagocytosis rate is low enough to allow for persistent parasitaemia. In these chronic infections, splenic retention of uRBCs and iRBCs stimulates a ten-fold increase in the splenic macrophage population, from an initial value of 1.2 × 10^9^ up to 13.1 × 10^9^ for Pf and 12.9 × 10^9^ for Pv. With our chosen iRBC phagocytosis rate (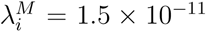 per macrophage), this corresponds to hourly phagocytosis probabilities of 0.178 for Pf and 0.176 for Pv. These phagocytosis rates are insufficient to control and overcome accumulation of parasites in the spleen.

The removal of uRBCs from circulation due to splenic retention was shown to be tens of times larger than the uRBC loss due to parasitisation in chronic infections, with the ratio of circulating uRBCs lost to splenic retention per circulating uRBC lost to parasitisation estimated to be 17:1 for Pf (5.5% due to parasitisation) and 82:1 for Pv (1.2% due to parasitisation). These ratios are larger than previous estimates for uRBC loss in acute clinical infections. These studies estimated that circulating RBC loss due to parasitisation, as a proportion of overall circulating RBC loss, was 10.5% (a ratio of 1:8.5) for acute Pf malaria in non-immune neurosyphilis patients (Jakeman et al., 1999); 7.9% (a ratio of 1:11.7) for acute clinical Pf malaria (Price et al., 2001); 2.9% (a ratio of 1:33.5) for acute Pv malaria in non-immune neurosyphilis patients (Collins et al., 2003); and 0.015% in Pf and Pv induced blood stage malaria volunteer infection studies (Woolley et al., 2021). In our model simulations this proportion differs by several orders of magnitude between the initial and chronic stages of asymptomatic Pf infections (0.01% and 5.5%, respectively) and asymptomatic Pv infections (0.02% and 1.2%, respectively). While our model was not calibrated to characterise acute symptomatic infections, the very early stage proportions in our model are consistent with the 0.015% reported in pre-symptomatic stages of Pf and Pv infection as reported by Woolley et al. (2021). This finding is consistent with major variations in the proportion of uRBC removed by the spleen versus parasitisation across the course of infection.

In order for our model to reproduce the highest levels of uRBC retention observed in the asymptomatic Papuan data (≥ 10%), we had to substantially decrease the iRBC removal rate. This resulted in greater parasitaemia and increased uRBC splenic retention, resulting in extremely low haemoglobin levels (5–7 g/dL). In a follow-up study of splenic RBC retention in 37 adult Papuans who underwent splenectomy for trauma or hyperreactive splenomegaly, Kho et al. (2023) reported that splenic RBC load was negatively correlated with haemoglobin levels, reflecting lower numbers of circulating uRBCs in patients with splenomegaly. Patients with the highest levels of RBC retention by spleen-mimetic filtration in vivo (≥ 10%) had haemoglobin levels of ≈ 6 g/dL, consistent with the results obtained from our model.

Primary limitations of this model include the simplicity of the splenic retention rates, which does not explicitly account for the impact of RBC congestion in the spleen that may affect the ability for RBCs to flow through the spleen and return to the circulation, as suggested by the rapid retention of heated RBCs in the presence of splenomegaly (Looareesuwan et al., 1987). Our model also did not account for erythropoiesis events occurring in the spleen during malaria (Kho, 2024), however its contribution to the size of splenic uRBC populations is likely negligible. Finally, the model does not explain the mechanics by which the splenic phagocytosis capacity might be dysfunctional or simply overwhelmed in immune or semi-immune individuals such as the Papuan patients. A recent observation in sickle cell disease has shown that the splenic density of macrophages was normal but not adapted to the intensity of congestion Sissoko et al. (2024).

The primary limitation of our analysis is the lack of longitudinal RBC data for asymptomatic individuals, which meant that we were only able to calibrate the model against the reported cross-sectional data. For this reason we kept parameter values constant, rather than allowing them to vary over time. Accordingly, the model results for the initial stages of infection do not necessarily reflect the true progression from initial exposure to chronic infection, since many of these parameters will vary over the course of an infection. We also used identical values for Pf and Pv infections where possible, and the sensitivity analysis demonstrated that the model results were not sensitive to moderate adjustments to our reference values. However, parameters such as the parasite multiplication factor likely differ for Pf and Pv, given differing abundance of target cells.

In summary, we have established the first modelling framework to study RBC dynamics in Pf and Pv infections that includes the spleen as a separate compartment where iRBCs and uRBCs can be retained and phagocytosed, consistent with recent knowledge advancements in malaria biology involving the spleen as a hidden compartment. Our findings are consistent with experimental data and suggest that the spleen can act as a reservoir for iRBCs and sustain chronic asymptomatic malaria infections, when the iRBC retention rate exceeds the phagocytosis capacity of the splenic macrophage population in the red-pulp. By quantifying RBC loss in this model, we also show that the vast majority of RBCs lost in chronic malaria are lost to splenic retention of uRBCs and not to obligatory destruction of iRBCs.

## 5 Availability of materials

The model code, input data sets, and generated outputs are available in a public repository: https://gitlab.unimelb.edu.au/rgmoss/malaria-spleen-rbc-loss. The model is implemented in R and provided as an R package (spleenrbc).

## Supporting information

Supplementary material: complete model description

## Data Availability

All data produced are available online at https://gitlab.unimelb.edu.au/rgmoss/malaria-spleen-rbc-loss

https://rgmoss.pages.gitlab.unimelb.edu.au/malaria-spleen-rbc-loss/

## Acknowledgements

J.A.S. is funded by an Australian NHMRC Leadership Investigator Grant (1196068). R.M., M.R. and D.J.P. are partly funded through Australian NHMRC CRE Grant 2024622 (Australian Centre of Research Excellence for Malaria Elimination) and Australian NHMRC Synergy Grant 2018654. S.K. was supported by an Australian NHMRC Emerging Leadership Investigator Grant (2025376) and Australian NHMRC Ideas Grant 2019153. N.M.A. is supported by Australian NHMRC Program Grant 1037304. B.B is funded by an Australian NHMRC Emerging Leadership Investigator Grant (2016792). P.A.B. is funded by Agence Nationale de la Recherche Grant ANR-24-CE17-2900-01 (“HepCMal”).

## A Mathematical model

A complete model description is provided in the supplementary materials, and is also published online^1^. See the Evidence for parameter values^2^ vignette (published online) for further details of how parameter values were chosen.

Unparasitised RBCs can be lost from circulation due to parasitisation, but also due to increased uRBC retention in the spleen due to the retention of iRBCs in the spleen. As shown in Figure A1, both causes of uRBC loss increase substantially from the inintial stage to chronic infection, but increased splenic uRBC retention is consistently the primary cause for uRBC loss from circulation.

Figure A2 compares the sensitivity analysis steady-state results (150 days from the initial infection) to the data collected from the splenectomised patients. The model can produce a similar distribution of iRBC counts in the circulation and spleen, but exhibits substantially less variation in uRBC counts than observed in the splenectomised patients. This may be due (at least in part) to our use of a fixed initial model state, which was informed by data collected in a cross-sectional household survey (Pava et al., 2016).

We also present uncertainty intervals for red blood cell counts and ratios of interest, reported over a period of 150 days from the initial infection. Figure A3 shows uncertainty intervals for uRBC and iRBC counts in the circulation and spleen. Figure A4 shows uncertainty intervals for the iRBC biomass ratio (spleen versus circulation). Figure A5 shows uncertainty intervals for the uRBC loss ratio (splenic retention versus parasitisation).

**Figure A1:**
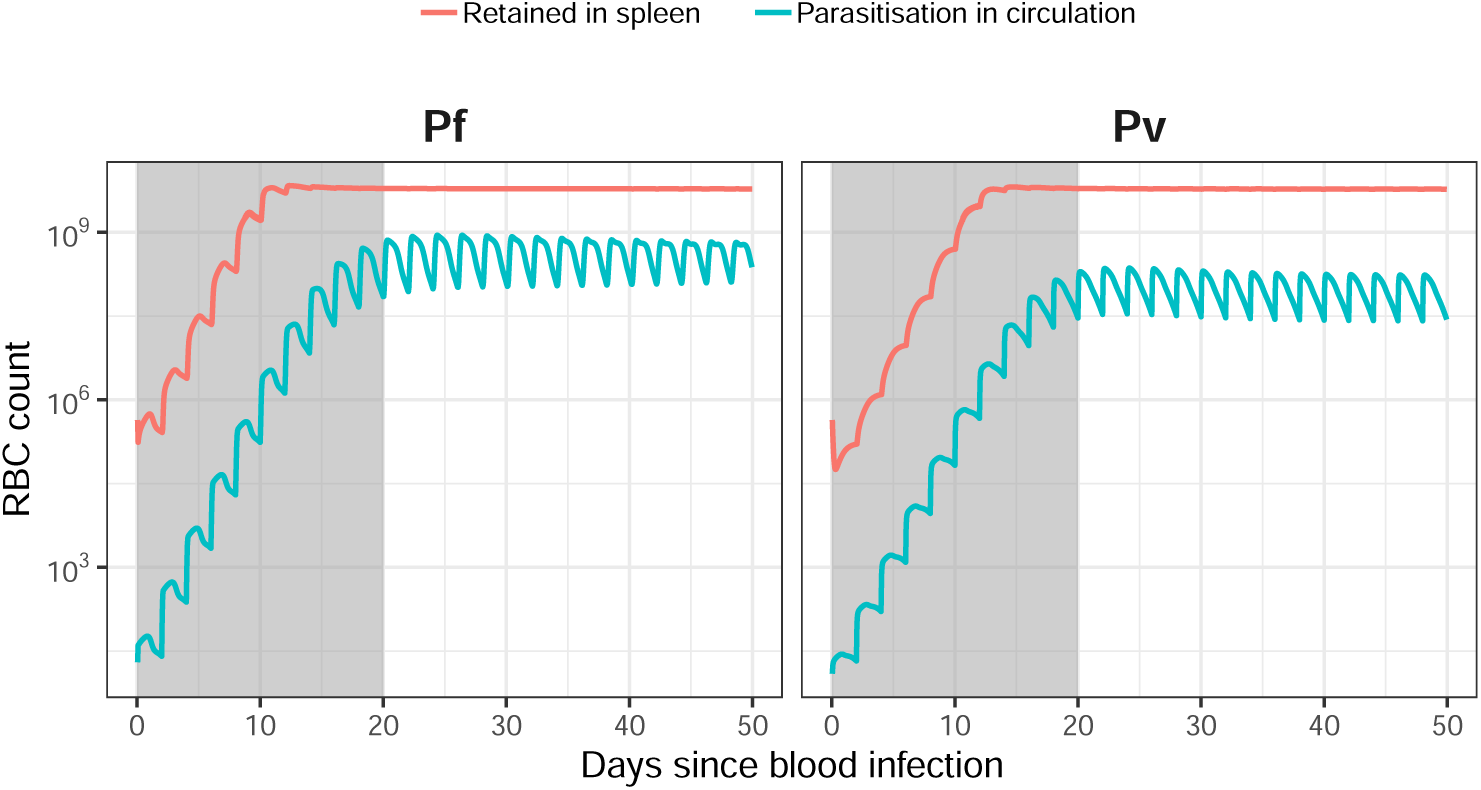
Unparasitised RBC loss from circulation due to malaria, as a consequence of (a) increased uRBC retention in the spleen (red lines); and (b) infection from parasites released in the circulation and spleen (blue lines). Increase splenic retention of uRBCs is substantially higher than parasitisation of circulating uRBCs at all stages of both Pf and Pv infections. Shaded intervals indicate the acute infection phase (i.e., the transition from initial infection to chronic infection), which the model dynamics do not capture.

**Figure A2:**
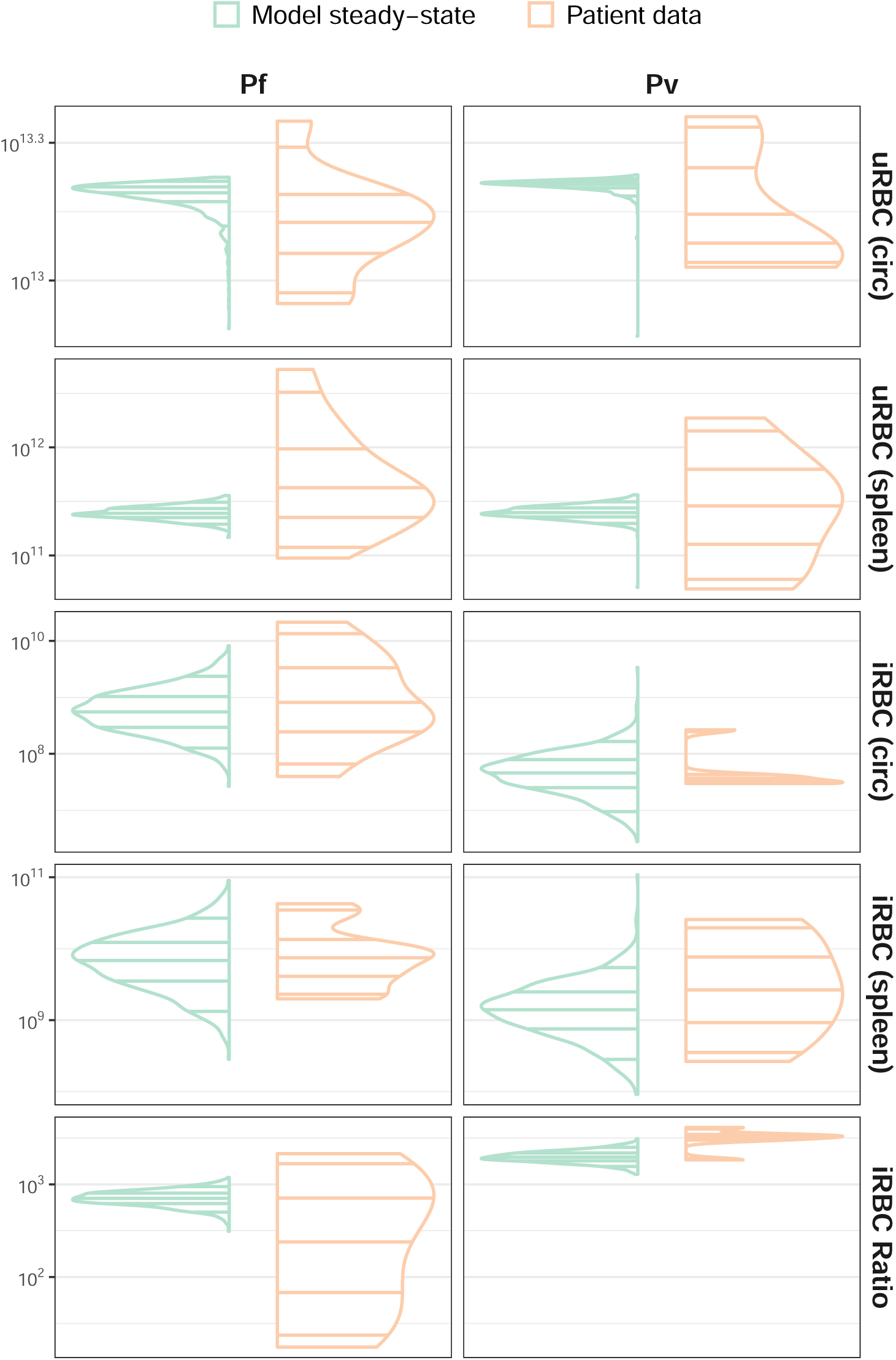
Sensitivity analysis results: the steady-state distributions for uRBC and iRBC counts in the circulation and spleen, and the ratio of iRBC biomass in the spleen to the iRBC biomass in the circulation, as obtained at 150 days from the initial infection (green). Corresponding distributions in the splenectomised patients (Pf: n=9; Pv: n=6) are shown on the right of each plot for comparison (orange).

**Figure A3:**
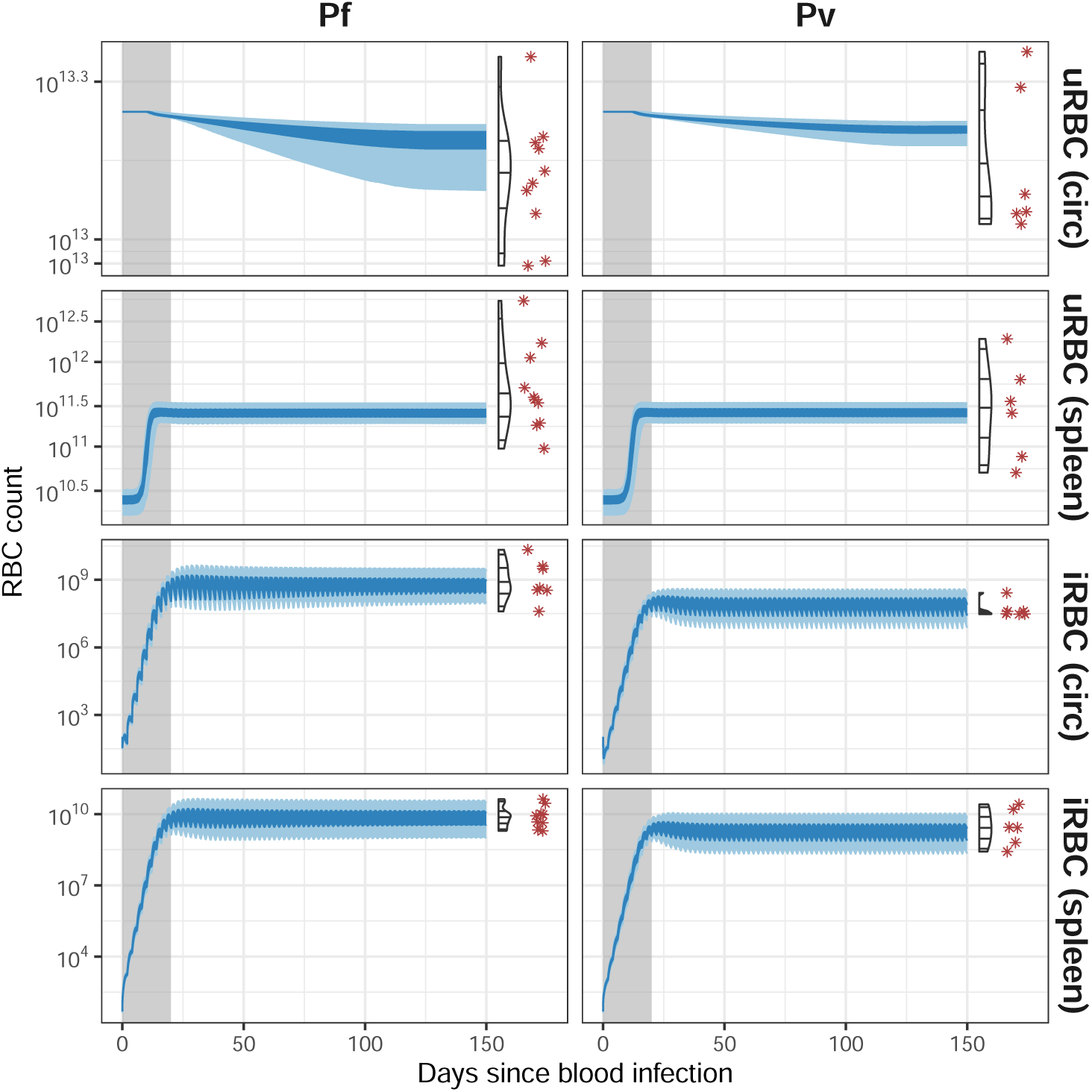
Sensitivity analysis results: uRBC and iRBC counts in the circulation and spleen, shown as 50% (dark blue shading) and 95% (light blue shading) uncertainty intervals, over a period of 150 days from the initial infection. Corresponding data in the splenectomised patients (Pf: n=9; Pv: n=6) are shown on the right of each plot for comparison (violin plots for distribution and red asterisks for observed data points). Shaded intervals indicate the acute infection phase (i.e., the transition from initial infection to chronic infection), which the model dynamics do not capture.

**Figure A4:**
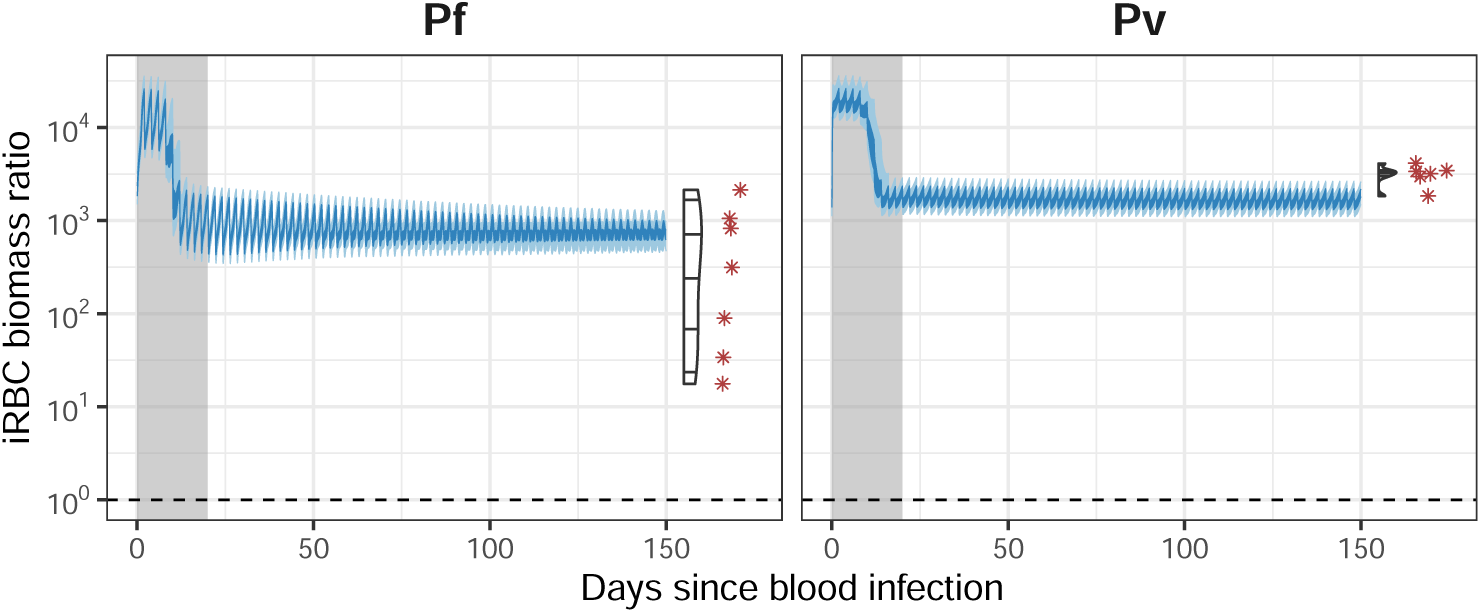
Sensitivity analysis results: the ratio of iRBC biomass in the spleen to the iRBC biomass in the circulation, shown as 50% (dark blue shading) and 95% (light blue shading) uncertainty intervals, over a period of 150 days from the initial infection. Corresponding data in the splenectomised patients (Pf: n=9; Pv: n=6) are shown on the right of each plot for comparison (violin plots for distribution and red asterisks for observed data points). Shaded intervals indicate the acute infection phase (i.e., the transition from initial infection to chronic infection), which the model dynamics do not capture.

**Figure A5:**
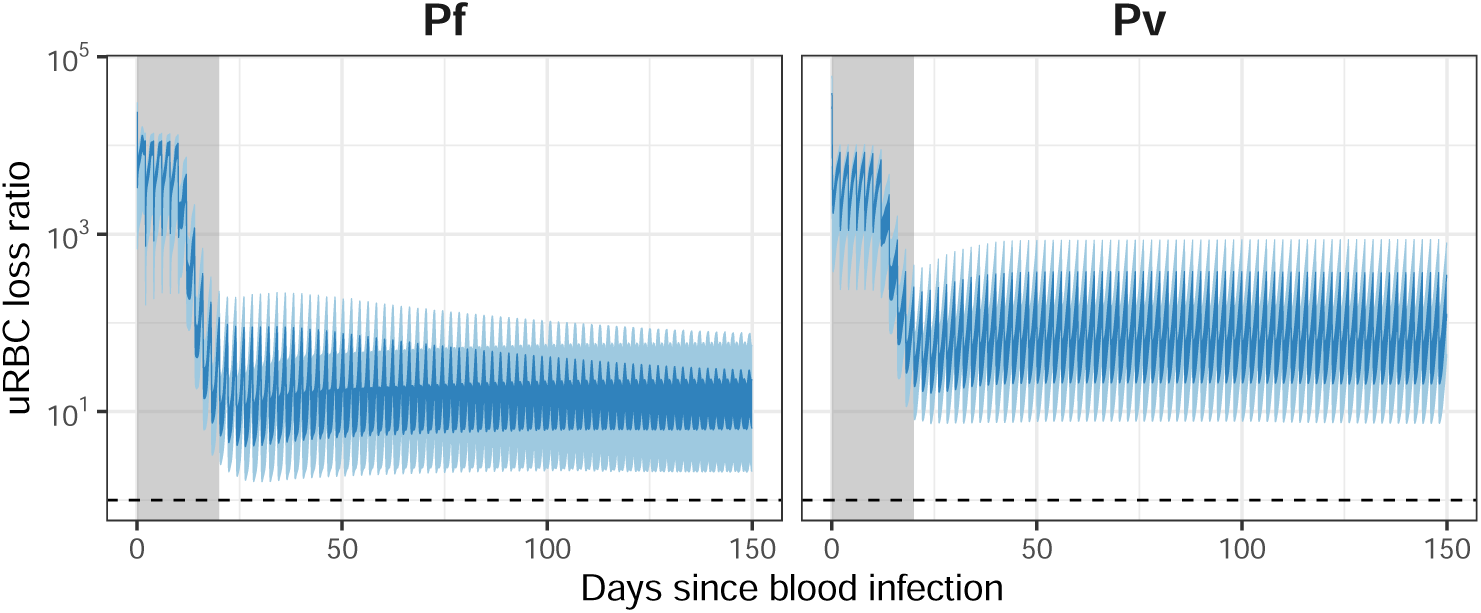
Sensitivity analysis results: the ratio of uRBC loss from circulation due to retention in the spleen to the uRBC loss from circulation due to infection by malaria parasites, shown as 50% (dark blue shading) and 95% (light blue shading) uncertainty intervals, over a period of 150 days from the initial infection. Shaded intervals indicate the acute infection phase (i.e., the transition from initial infection to chronic infection), which the model dynamics do not capture.

## Footnotes

1 https://rgmoss.pages.gitlab.unimelb.edu.au/malaria-spleen-rbc-loss/articles/model-description.html

2 https://rgmoss.pages.gitlab.unimelb.edu.au/malaria-spleen-rbc-loss/articles/evidence-for-parameter-values.html

