## Supplementary material: complete model description for "The role of the spleen in red blood cell loss caused by malaria: a mathematical model"

#### Contents


**S1 Compartments and cell populations** **S2**

**S2 Homeostasis and initial state** **S3**

**S3 RBC production in the bone marrow** **S7**

**S4 RBC release from the bone marrow** **S9**

**S5 Uninfected RBC removal from circulation** **S11**

**S6 Uninfected RBC return to circulation** **S13**

**S7 RBC infection** **S14**

**S8 Infected RBC removal from circulation** **S16**

**S9 Infected RBC return to circulation** **S19**

**S10 Infected RBC sequestration** **S21**

**S11 RBC destruction in the spleen** **S23**

**S12 Differences between Pf and Pv** **S24**

**S13 Final RBC equations** **S25**

**S14 Baseline outputs: no infection** **S26**

**S15 Baseline outputs: Pf** **S28**

**S16 Baseline outputs: Pv** **S31**

**S17 Model parameters** **S34**

References **S35**

### S1 Compartments and cell populations

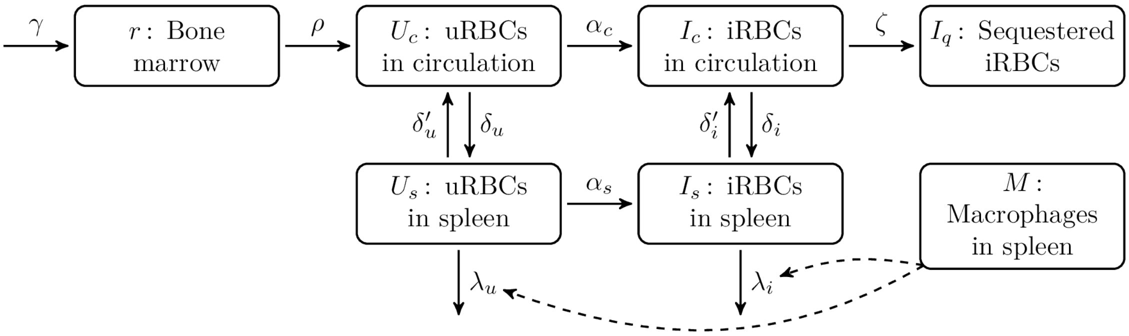


Figure S1: An overview of the model structure.

Table S1: The cell populations in the model, with respect to age $a$ and time $t$ where appropriate.

| Symbol | Compartment | Cell type | Lifespan (days) |
|---|---|---|---|
| $r(a,t)$ | Bone marrow | Normoblasts and reticulocytes | $T_R = 3.5$ |
| $R_c(a,t)$ | Circulation | Uninfected reticulocytes | $T_M = 4.5$ |
| $N_c(a,t)$ | Circulation | Uninfected normocytes | $T_N = 115.5$ |
| $U_c(a,t)$ | Circulation | Uninfected RBCs | $T_U = 120$ |
| $I_c(a,t)$ | Circulation | Infected RBCs | $T_I = 2$ |
| $I_q(a,t)$ | Microvasculature | Infected RBCs | $T_I = 2$ |
| $U_s(a,t)$ | Spleen | Uninfected RBCs | $T_U = 120$ |
| $I_s(a,t)$ | Spleen | Infected RBCs | $T_I = 2$ |
| $M(t)$ | Spleen | Macrophages | — |

For convenience, we also define terms for the total RBC populations in the circulation and the spleen:

$$\mathbf{U_c}(t) \equiv \sum_a U_c(a,t) \tag{1}$$

$$\mathbf{U_s}(t) \equiv \sum_a U_s(a,t) \tag{2}$$

$$\mathbf{I_c}(t) \equiv \sum_a I_c(a,t) \tag{3}$$

$$\mathbf{I_s}(t) \equiv \sum_a I_s(a,t) \tag{4}$$

#### S2 Homeostasis and initial state

Some model parameters are defined relative to the steady-state (homeostasis) in the absence of a malaria infection, where the circulating reticulocyte and normocyte populations — $R_c(a,t)$ and $N_c(a,t)$, respectively — are equal to the steady-state RBC population $U_{ss}$. The value of $U_{ss}$ is the median RBC count (in the circulation) for patients with no fever (`fever24 == 0`) or malaria infection (`Species == 0`).

```
utils::data("rbc_steady_state", package = "spleenrbc")
rbc_steady_state
#> [1] 1.75197e+13
```

We use a root-finding method to solve the following conservation equation for the normoblast production rate $\gamma$ *in the absence of a malaria infection*:

$$U_{ss} \approx \sum_a N_c(a) + \sum_a R_c(a) : r(1) = \gamma \quad (5)$$

```
p <- baseline_parameters("Pf")
steady_state <- retic_steady_state(p)
print(steady_state$gamma)
#> [1] 7202963644
print(sum(steady_state$r_a))
#> [1] 580623948414
print(sum(steady_state$R_a))
#> [1] 181144300035
print(sum(steady_state$N_a))
#> [1] 1.733856e+13
print(sum(steady_state$Ur_a))
#> [1] 23347264546
```

Note that **only a very small fraction** of the RBCs are retained in the spleen at homeostasis:

```
rbc_spleen <- sum(steady_state$Ur_a)
rbc_circ <- sum(c(p$R_a_ss, p$N_a_ss))
pcnt_in_spleen <- 100 * rbc_spleen / (rbc_spleen + rbc_circ)
cat("Retained RBCs:", sprintf("%0.2f%%", pcnt_in_spleen), "\n")
#> Retained RBCs: 0.13%
```

Table S2: The steady-state model parameters. Note that some are age-dependent.

| Symbol | Description | Baseline value |
|---|---|---|
| $U_{ss}$ | RBC population | $1.75197 \times 10^{13}$ |
| $r_{ss}(a)$ | Reticulocyte population (bone marrow) | $5.8062395 \times 10^{11}$ |
| $R_{ss}(a)$ | Reticulocyte population (circulation) | $1.811443 \times 10^{11}$ |
| $N_{ss}(a)$ | Normocyte population (circulation) | $1.7338556 \times 10^{13}$ |

| Symbol | Description | Baseline value |
|---|---|---|
| $U_{s,ss}(a)$ | RBC population (spleen) | $2.3347265 \times 10^{10}$ |
| $M_0$ | Initial macrophage population | $1.2 \times 10^9$ |
| $I_0$ | Initial infected RBC population | 100 |

```
s0 <- initial_spleenrbc_state(p)
```

These steady-state parameters define the initial (uninfected) cell populations:

$$r(a, t = 0) = r_{ss}(a) \tag{6}$$
$$R_c(a, t = 0) = R_{ss}(a) \tag{7}$$
$$N_c(a, t = 0) = N_{ss}(a) \tag{8}$$
$$U_s(a, t = 0) = U_{s,ss}(a) \tag{9}$$
$$M(t = 0) = M_0 \tag{10}$$

We start with a small number $I_0$ of infected RBCs in the circulation, with a small bias towards middle-aged cells (using a truncated normal distribution), and some infected RBCs in the spleen and the microvasculature:

$$I_c(a, t = 0) = I_0 \cdot f_X(a) : a \sim \mathcal{N}(\mu = 20, \sigma = 30) \tag{11}$$
$$I_q(a, t = 0) = I_c(a, t = 0) \cdot (1 - \exp[-\zeta]) \tag{12}$$
$$I_s(a, t = 0) = I_c(a, t = 0) \cdot (1 - \exp[-\delta_i]) \cdot \exp(-\lambda_i) \tag{13}$$

The initial cell populations for the baseline parameter values are shown in Figures S2, S3, and S4.

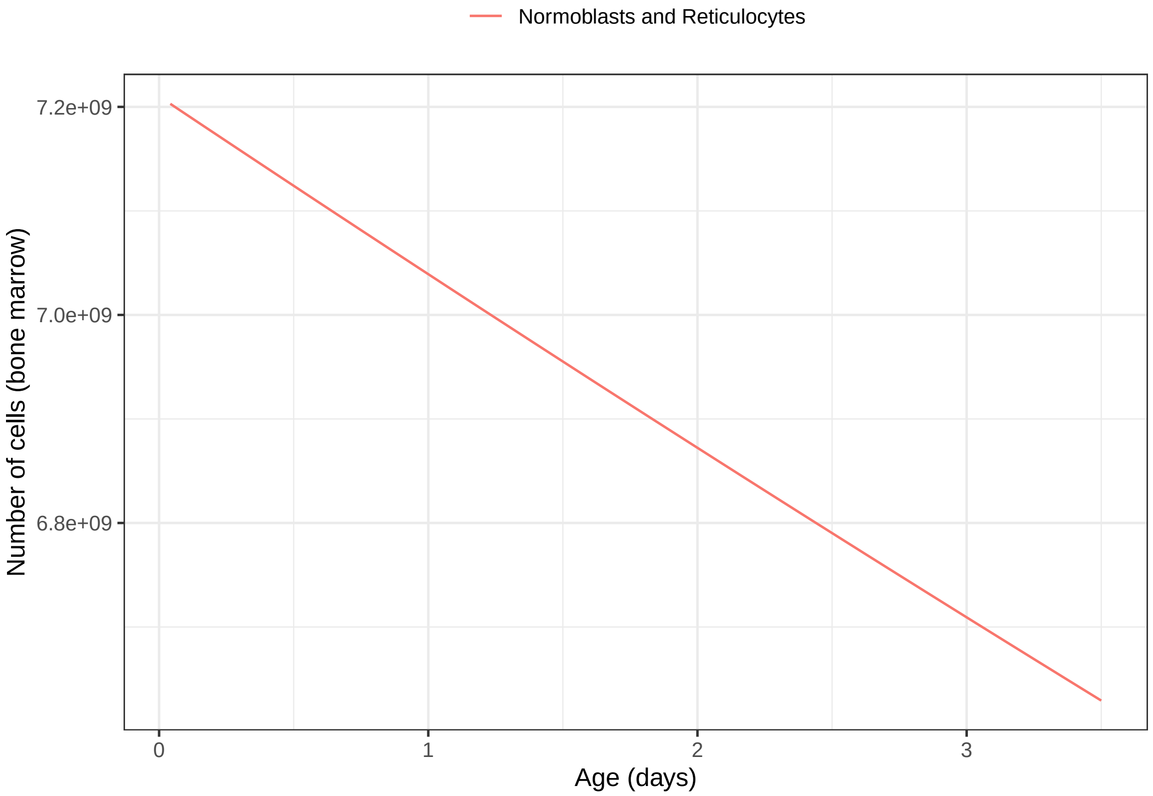


Figure S2: Initial cell populations in the bone marrow.

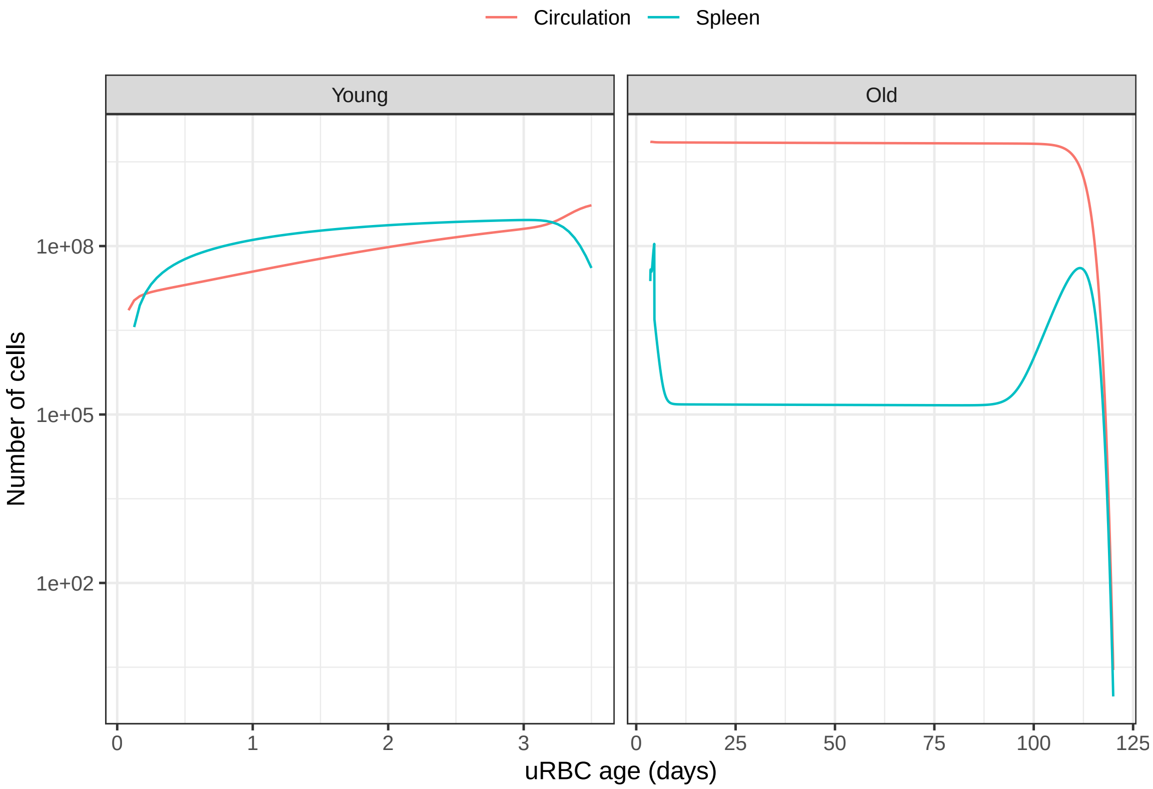


Figure S3: Initial uninfected RBC populations.

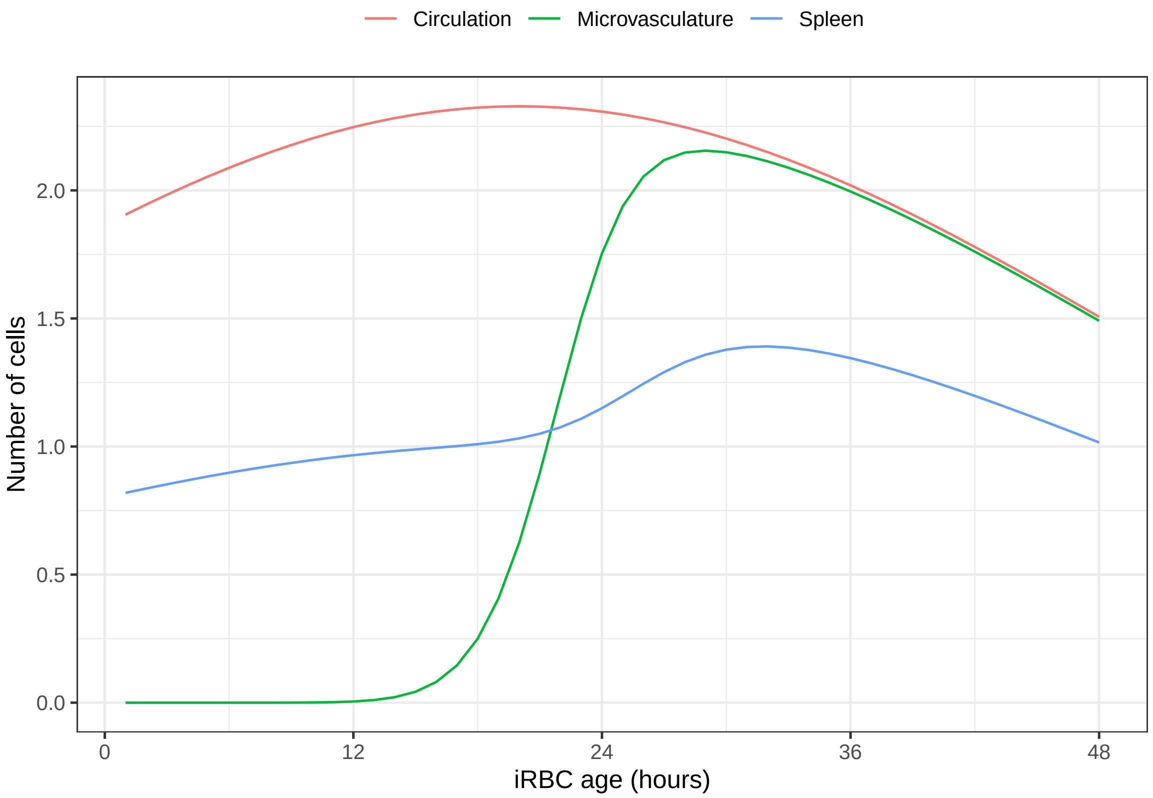


Figure S4: Initial infected RBC populations.

#### S3 RBC production in the bone marrow

The normoblast production rate depends on the population of circulating uninfected RBCs $\mathbf{U_c}(t)$, the steady-state uninfected RBC population $U_{ss}$, the steady-state normoblast production rate $\gamma$, and the threshold parameter $U_c^l$:

$$U_l = U_c^l \cdot U_{ss} \tag{14}$$

$$\text{eryth}(t) = \gamma \cdot \left[1 + \frac{f_{\text{max}} - 1}{2} \cdot \left[1 - \tanh(e_{\text{sl}} \cdot \frac{\mathbf{U_c}(t) - U_l}{U_l})\right]\right] \tag{15}$$

Table S3: Model parameters for erythropoiesis.

| Symbol | Description | Baseline value |
|---|---|---|
| $\gamma$ | Steady-state normoblast production | $7.2029636 \times 10^9$ |
| $U_c^l$ | Threshold parameter | 0.33 |
| $f_{\text{max}}$ | Scaling parameter | 10 |
| $e_{\text{sl}}$ | Slope parameter | 16 |

Our chosen value of $f_{\text{max}}$ is consistent with a previous modelling study (Watson et al., 2017), which noted that "in extreme anaemia [RBC production] can be increased fivefold or more", and produced model fits where RBC production was increased almost ten-fold.

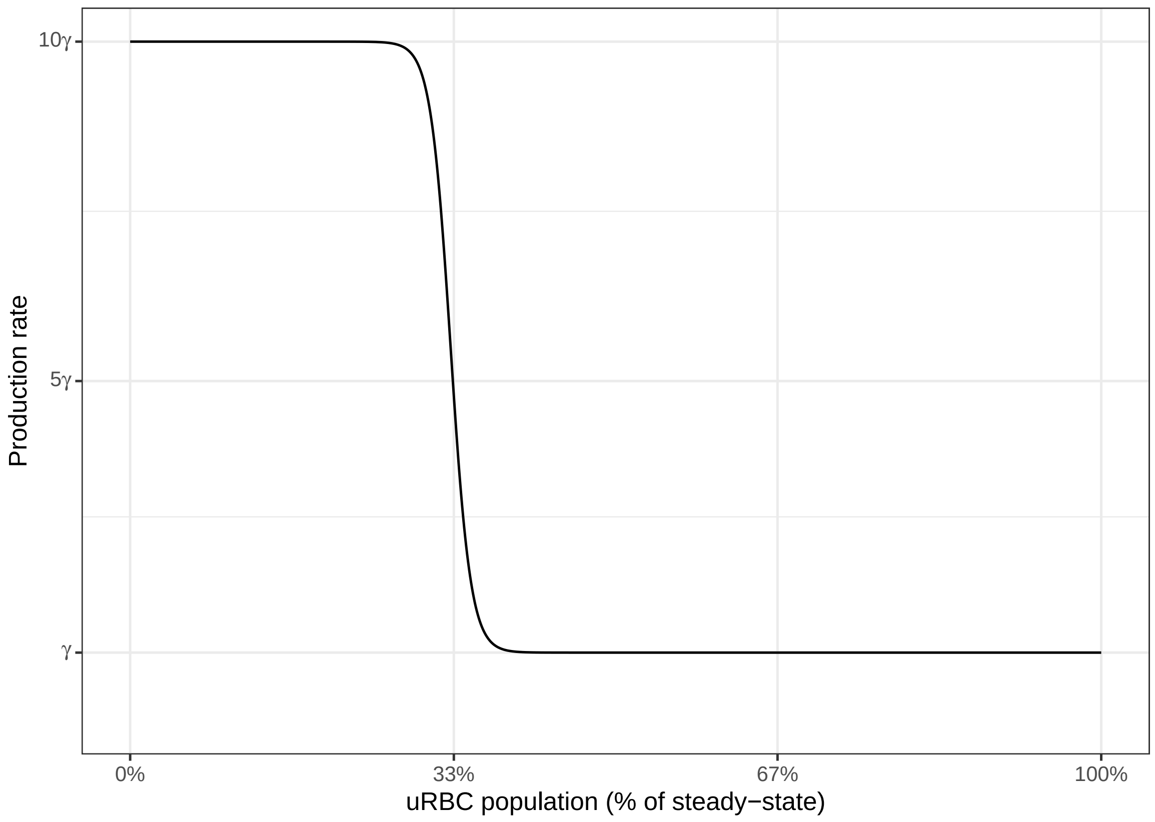


Figure S5: The erythropoiesis rate varies from $\gamma$ to $f_{\max} \cdot \gamma$.

#### S4 RBC release from the bone marrow

The normoblast and reticulocyte populations in the bone marrow age over time, and the reticulocytes move into the circulation at the age-specific rate $\rho(a, t)$:

$$r(a,t) = \begin{cases} \text{eryth}(t) & \text{for } a = 1 \\ r(a-1, t-1) \cdot \exp[-\rho] & \text{for } 1 < a \leq T_R \end{cases} \tag{16}$$

$$\rho(a,t) = \begin{cases} 10 \cdot \min(20, \exp[\kappa \cdot (U_{ss} - \mathbf{U_c}(t))]) & \text{for } a \geq t_r \\ \rho_o \cdot \min(20, \exp[\kappa \cdot (U_{ss} - \mathbf{U_c}(t))]) & \text{for } a < t_r \end{cases} \tag{17}$$

$$t_r = \begin{cases} T_R & \text{when } \mathbf{U_c}(t) > U_{ss} \\ T_R^{\min} + (T_R - T_R^{\min}) \cdot \left(1 + \exp\left[-\rho_s \cdot \left(\frac{\mathbf{U_c}(t)}{U_{ss}} - \rho_i\right)\right]\right)^{-1} & \text{when } \mathbf{U_c}(t) \leq U_{ss} \end{cases} \tag{18}$$

Table S4: Model parameters for bone marrow cell equations.

| Symbol | Description | Baseline value |
|---|---|---|
| $\rho_0$ | Minimum release rate | 0.001 |
| $\rho_s$ | Slope parameter | 10 |
| $\rho_i$ | Inflection parameter | 0.5 |
| $\kappa$ | Scaling factor | $10^{-9}$ |
| $T_R^{\min}$ | Minimum retention time | 24 hours |
| $T_R$ | Reticulocyte release time | 84 hours |

These parameters were calibrated using data from Koepke and Koepke (1986), as shown in Figure S6.

We denote the number of reticulocytes released from the bone marrow in a time-step as $r_{\to c}$:

$$r_{\to c}(a,t) = r(a-1, t-1) \cdot (1 - \exp[-\rho]) \tag{19}$$

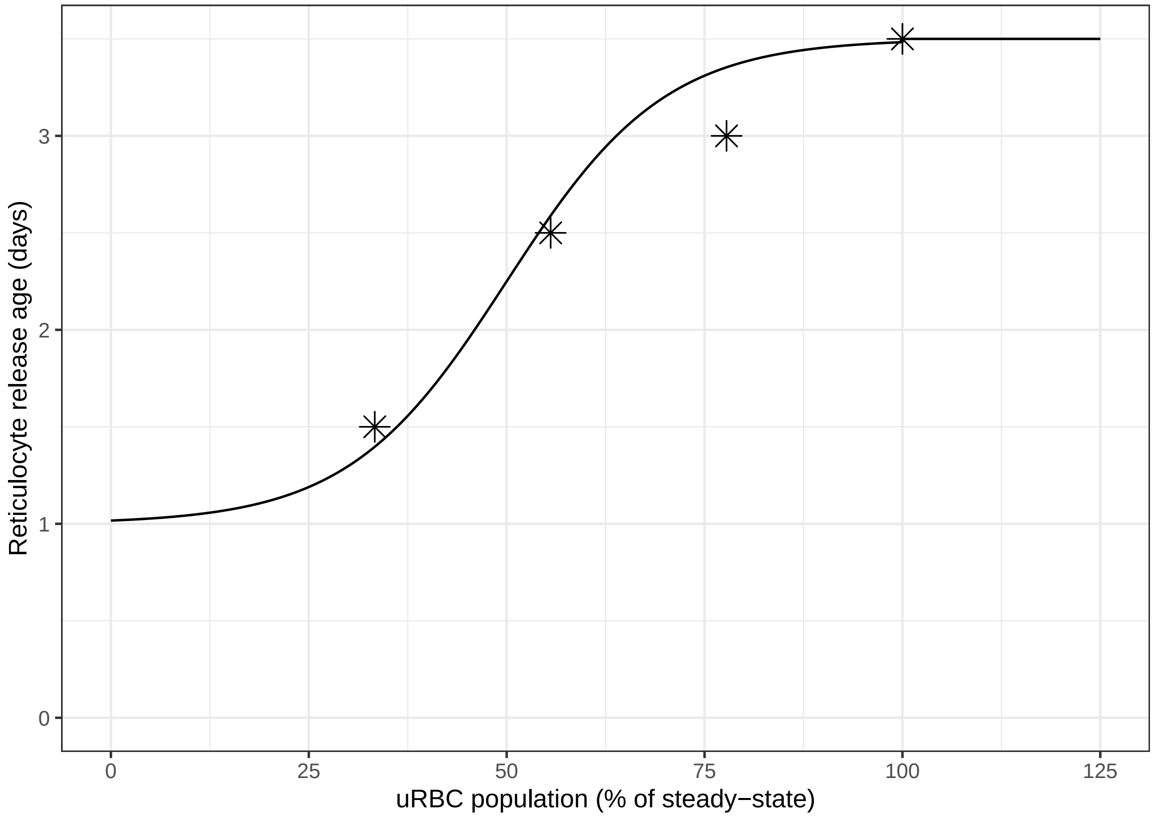


Figure S6: The minimum age at which reticulocytes are released from the bone marrow. Point estimates are from Figure 2 of Koepke and Koepke (1986).

#### S5 Uninfected RBC removal from circulation

The removal rate $\delta_u$ of uninfected RBCs from the circulation into the spleen depends on a number of offset, slope, and scaling parameters:

$$\delta_u(a,t) = F_U(t) \cdot \left( \delta_U^A \exp[-k_1 \cdot (a-1)] + \nu + \delta_U^{\min} + (\delta_U^{\max} - \delta_U^{\min}) \cdot \frac{(a-1)^{\delta_U^g}}{(a-1)^{\delta_U^g} + (\delta_U^{c50})^{\delta_U^g}} \right) \tag{20}$$

$$F_U(t) = 1 + k_\nu^U \cdot \frac{\mathbf{I_c}(t-1)^{g_d^U}}{\mathbf{I_c}(t-1)^{g_d^U} + \left(\delta_{50}^U \cdot [\mathbf{I_c}(t-1) + \mathbf{U_c}(t-1)]\right)^{g_d^U}} \tag{21}$$

$$k_1 = -\frac{\log\left(\frac{\delta_U^{\min}}{\delta_U^A}\right)}{23 \cdot 7} \tag{22}$$

| Symbol | Description | Baseline value |
|---|---|---|
| $\delta_U^A$ | Scaling parameter | 0.74303 |
| $\delta_U^{\min}$ | Scaling parameter | $2.16405 \times 10^{-5}$ |
| $\delta_U^{\max}$ | Scaling parameter | 1.206914 |
| $\delta_U^{c50}$ | Half-maximal age | 2954.306 |
| $\delta_U^g$ | Scaling parameter | 43.73335 |
| $\nu$ | Offset parameter | 0 |
| $k_\nu^U$ | Scaling parameter | 1 |
| $g_d^U$ | Slope parameter | 1 |
| $\delta_{50}^U$ | Scaling parameter | $10^{-7}$ |

We denote the number of uRBCs removed in a time-step as $U_{c\to s}$:

$$U_{c\to s}(a,t) = U_c(a-1, t-1) \cdot (1 - \exp[-\delta_u(a,t)]) \tag{23}$$

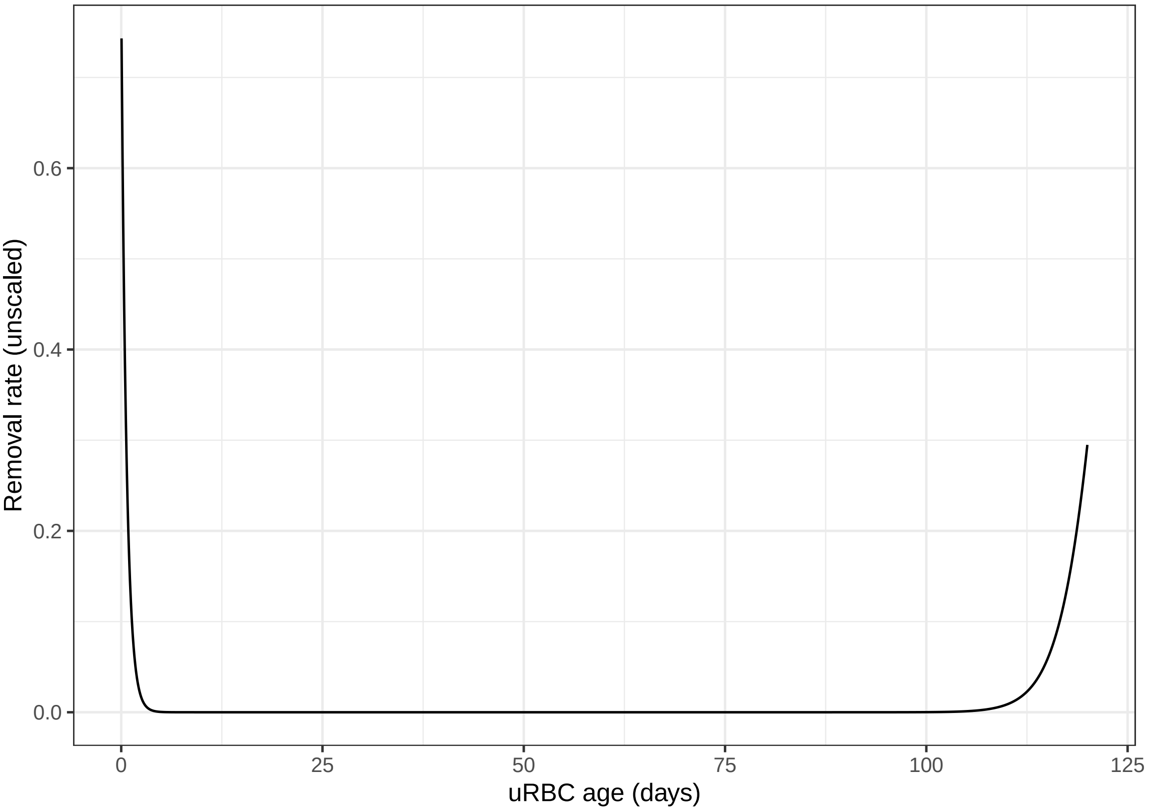


Figure S7: The uRBC removal rate $\delta_u(a, t)$ from the circulation into the spleen when the scaling factor $F_U = 1$.

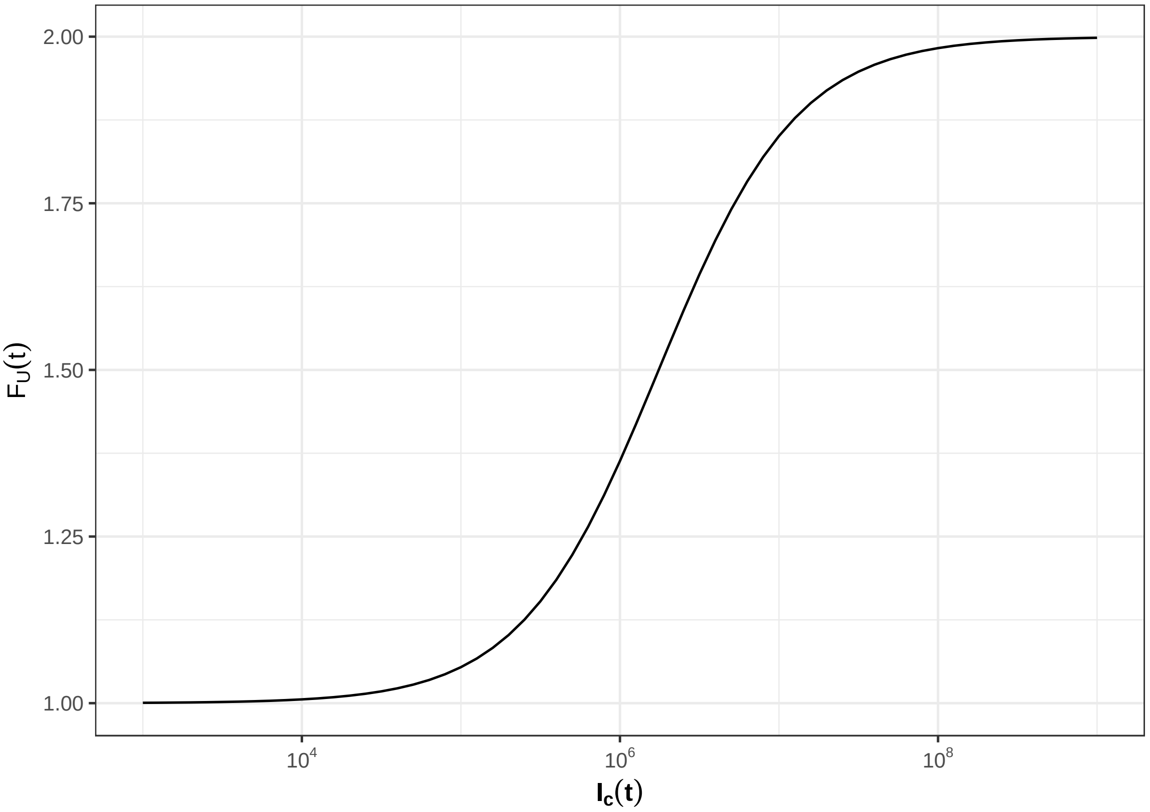


Figure S8: The fold increase in uRBC removal rate due to the presence of iRBCs in the circulation, shown for $\mathbf{U_c}(t) = U_{ss}$.

### S6 Uninfected RBC return to circulation

Uninfected RBCs in the spleen return to the circulation at rate $\delta'_u$, which is defined in terms of the log-normal probability density function $F_X$:

$$\delta'_u(a,t) = \text{mag} \cdot F_X(a) \tag{24}$$

$$\log(X) \sim \mathcal{N}(\mu = 24 \cdot \mu_U, \sigma = 24 \cdot \sigma_U) \tag{25}$$

| Symbol | Description | Baseline value |
|---|---|---|
| mag | Scaling parameter | 10 |
| $_{\mathrm{U}}$ | Scaling parameter | 3.65 |
| $_{\mathrm{U}}$ | Scaling parameter | 0.0025 |

We denote the number of uRBCs released in a time-step as $U_{s\to c}$:

$$U_{s\to c}(a,t) = U_s(a-1,t-1) \cdot \exp[-\lambda_u(a,t)] \cdot (1 - \exp[-\delta'_u(a)]) \tag{26}$$

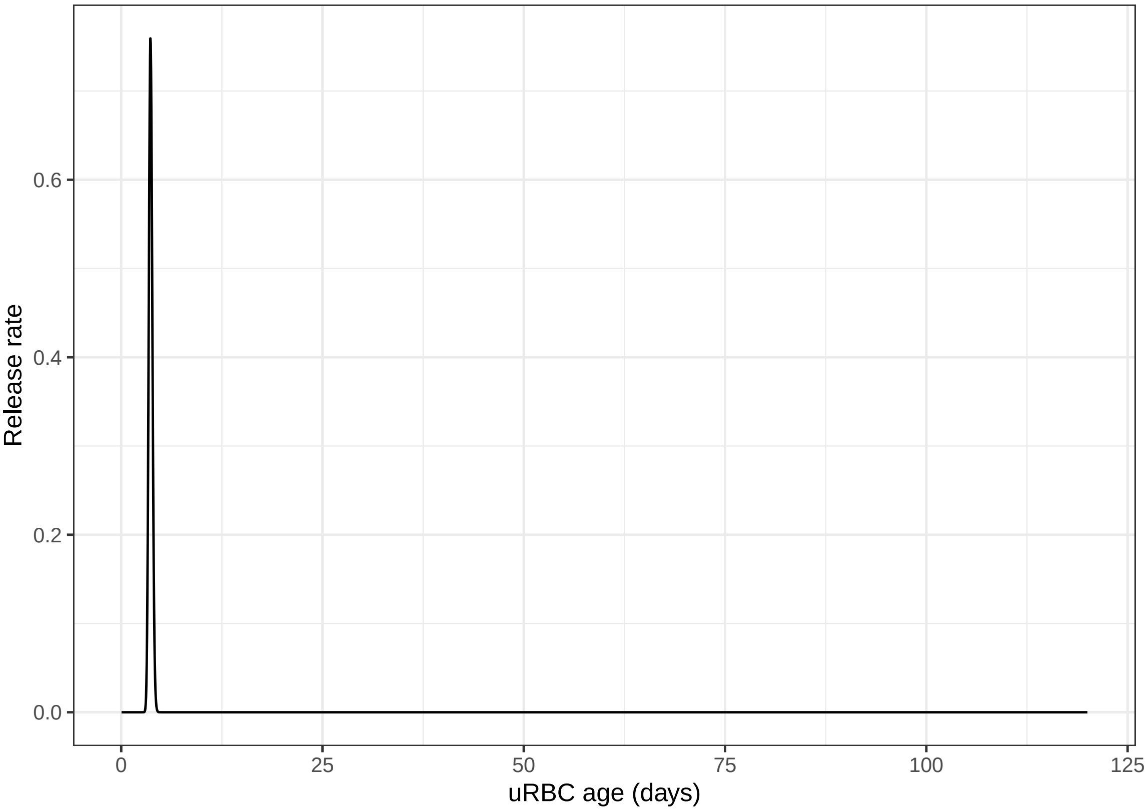


Figure S9: The uRBC release rate $\delta'_u(a,t)$ from the spleen into the circulation.

### S7 RBC infection

Uninfected RBCs are infected at rates $\alpha_c$ and $\alpha_s$ in the circulation and spleen, respectively, which depend on the age-specific merozoite preference $\beta$:

$$\alpha_c(a,t) = \text{PMF} \cdot \frac{\beta(a) \cdot U_c(a,t-1)}{\sum_{a'} [\beta(a') \cdot U_c(a',t-1)]} \tag{27}$$

$$\alpha_s(a,t) = \text{PMF} \cdot \frac{\beta(a) \cdot U_s(a,t-1)}{\sum_{a'} [\beta(a') \cdot U_s(a',t-1)]} \tag{28}$$

Merozoites are released when infected RBCs rupture at age $T_{\text{irbc}}$. Here we use the notation $\nabla_{x\to y}$ to define the number of uRBCs infected in location $y$ by merozoites released in location $x$:

$$\nabla_{c\to c}(a,t) = \alpha_c(a,t) \cdot [I_c^\nabla + I_q(T_{\text{irbc}}, t-1)] \tag{29}$$

$$\nabla_{s\to c}(a,t) = \alpha_c(a,t) \cdot \omega \cdot I_s^\nabla \tag{30}$$

$$\nabla_{s\to s}(a,t) = \alpha_s(a,t) \cdot (1-\omega) \cdot I_s^\nabla \tag{31}$$

These in turn are defined in terms of the iRBCs that remain in the circulation and spleen, respectively:

$$I_c^\nabla = I_c(T_{\text{irbc}}, t-1) \cdot \exp[-\delta_i] \cdot \exp[-\zeta] \tag{32}$$

$$I_s^\nabla = I_s(T_{\text{irbc}}, t-1) \cdot \exp[-\delta_i'] \cdot \exp[-\lambda_i] \tag{33}$$

We can then define the total number of infected RBCs in the circulation and in the spleen:

$$\nabla_c(t) = \sum_a \nabla_{c\to c}(a,t) + \sum_a \nabla_{s\to c}(a,t) \tag{34}$$

$$\nabla_s(t) = \sum_a \nabla_{s\to s}(a,t) \tag{35}$$

| Symbol | Description | Baseline value |
|---|---|---|
| PMF | Parasite multiplication factor | 8 |
| $\omega$ | Proportion of merozoites released in the spleen that infect circulating uRBCS | 0.1 |

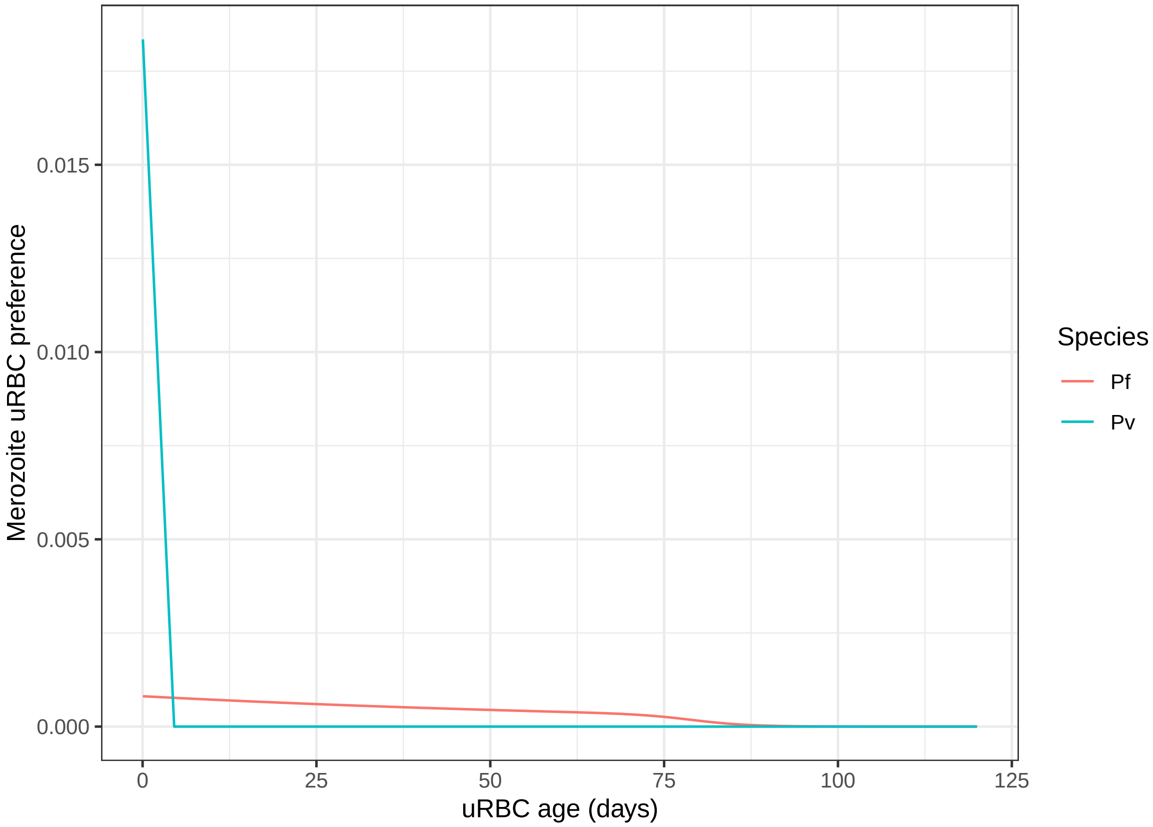


Figure S10: The age-dependent merozoite preference for uninfected RBCs.

#### S8 Infected RBC removal from circulation

The removal rate $\delta_i$ of infected RBCs from the circulation into the spleen depends on a number of offset, slope, and scaling parameters:

$$\delta_i(a,t) = F_I(t) \cdot \left[ \delta_{iR} + (\delta_{iS} - \delta_{iR}) \cdot \frac{(a-1)^{\delta_I^{sl}}}{(a-1)^{\delta_I^{sl}} + (\delta_I^{c50})^{\delta_I^{sl}}} \right] \tag{36}$$

$$F_I(t) = 1 + k_\nu^I \cdot \frac{\mathbf{I_c}(t-1)^{g_d^U}}{\mathbf{I_c}(t-1)^{g_d^U} + \left(\delta_{50}^I \cdot [\mathbf{I_c}(t-1) + \mathbf{U_c}(t-1)]\right)^{g_d^U}} \tag{37}$$

$$\delta_{iS} = \delta_{iR} \cdot k_{iS} \tag{38}$$

| Symbol | Description | Baseline value |
|---|---|---|
| $k_{iS}$ | Schizont scaling parameter | 2 |
| $\delta_{iR}$ | **Pf** ring removal rate | 0.562 |
| $\delta_{iS}$ | **Pf** schizont removal rate | 1.124 |
| $\delta_{iR}$ | **Pv** ring removal rate | 0.562 |
| $\delta_{iS}$ | **Pv** schizont removal rate | 1.124 |
| $\delta_I^{sl}$ | Slope parameter | 10 |
| $\delta_I^{c50}$ | Half-maximal age | 26 |
| $k_\nu^I$ | Scaling parameter | 3 |
| $g_d^U$ | Slope parameter | 1 |
| $\delta_{50}^I$ | Scaling parameter | $10^{-4}$ |

Safeukui et al. (2008) conducted in vitro experiments that showed 11% and 20% of Pf rings and schizonts, respectively, are retained in the spleen in every passage of the iRBCs through the spleen. Accordingly, we assume here that $\delta_{iS} \approx 2 \cdot \delta_{iR}$.

We denote the number of iRBCs removed in a time-step as $I_{c\to s}$:

$$I_{c\to s}(a,t) = I_c(a-1,t-1) \cdot (1 - \exp[-\delta_i(a,t)]) \tag{39}$$

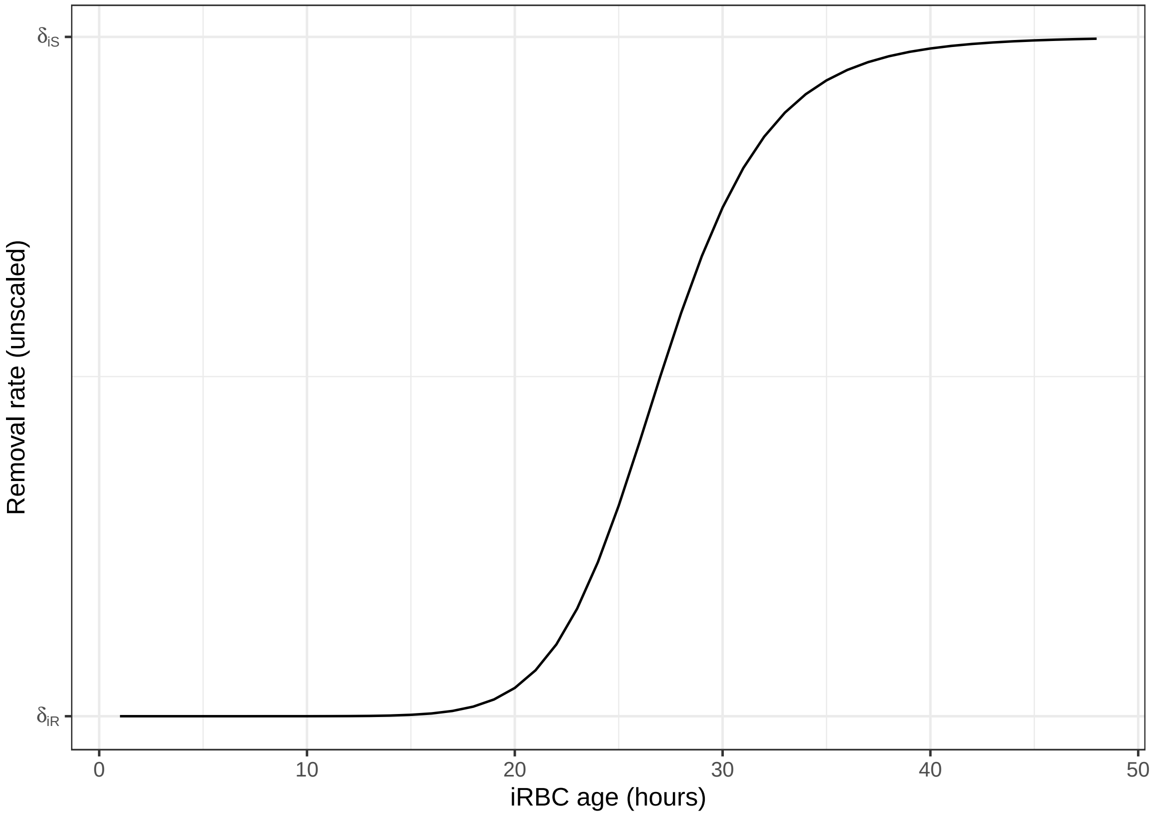


Figure S11: The iRBC removal rate $\delta_i(a, t)$ from the circulation into the spleen when the scaling factor $F_I = 1$.

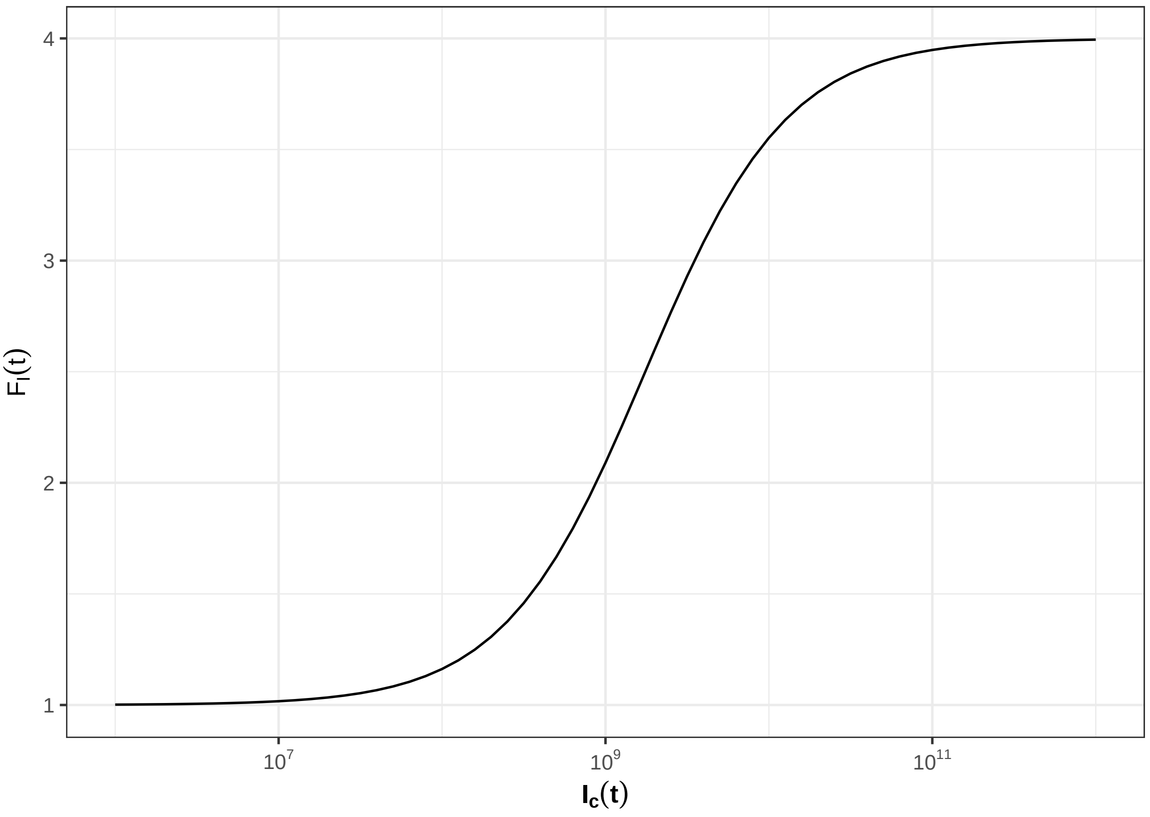


Figure S12: The fold increase in iRBC removal rate due to the presence of iRBCs in the circulation, shown for $\mathbf{U_c}(t) = U_{ss}$.

#### S9 Infected RBC return to circulation

Infected RBCs in the spleen return to the circulation at rate $\delta_i'$. We assume that ring-stage RBCs are more likely to return to the circulation than are schizont-stage RBCs, and that infected RBCs are returned at a much lower rate than they are removed from the circulation into the spleen.

$$\delta_i'(a) = \delta_{iR}' + (\delta_{iS}' - \delta_{iR}') \cdot \frac{(a-1)^{\delta_I^{sl}}}{(a-1)^{\delta_I^{sl}} + (\delta_I^{c50})^{\delta_I^{sl}}} \tag{40}$$

$$\delta_{iR}' = \delta_i R \cdot k_{iR} \tag{41}$$

$$\delta_{iS}' = \delta_i S \cdot k_{iS} \tag{42}$$

| Symbol | Description | Baseline value |
|---|---|---|
| $k_{iR}$ | Scaling parameter | 0.03 |
| $k_{iS}$ | Scaling parameter | 0.01 |
| $\delta_{iR}'$ | Ring iRBC return rate | 0.01686 |
| $\delta_{iS}'$ | Schizont iRBC return rate | 0.01124 |
| $\delta_I^{sl}$ | Slope parameter | 10 |
| $\delta_I^{c50}$ | Half-maximal age | 26 |

We denote the number of iRBCs released in a time-step as $I_{s\to c}$:

$$I_{s\to c}(a,t) = I_s(a-1,t-1) \cdot \exp[-\lambda_i(a,t)] \cdot (1 - \exp[-\delta_i'(a)]) \tag{43}$$

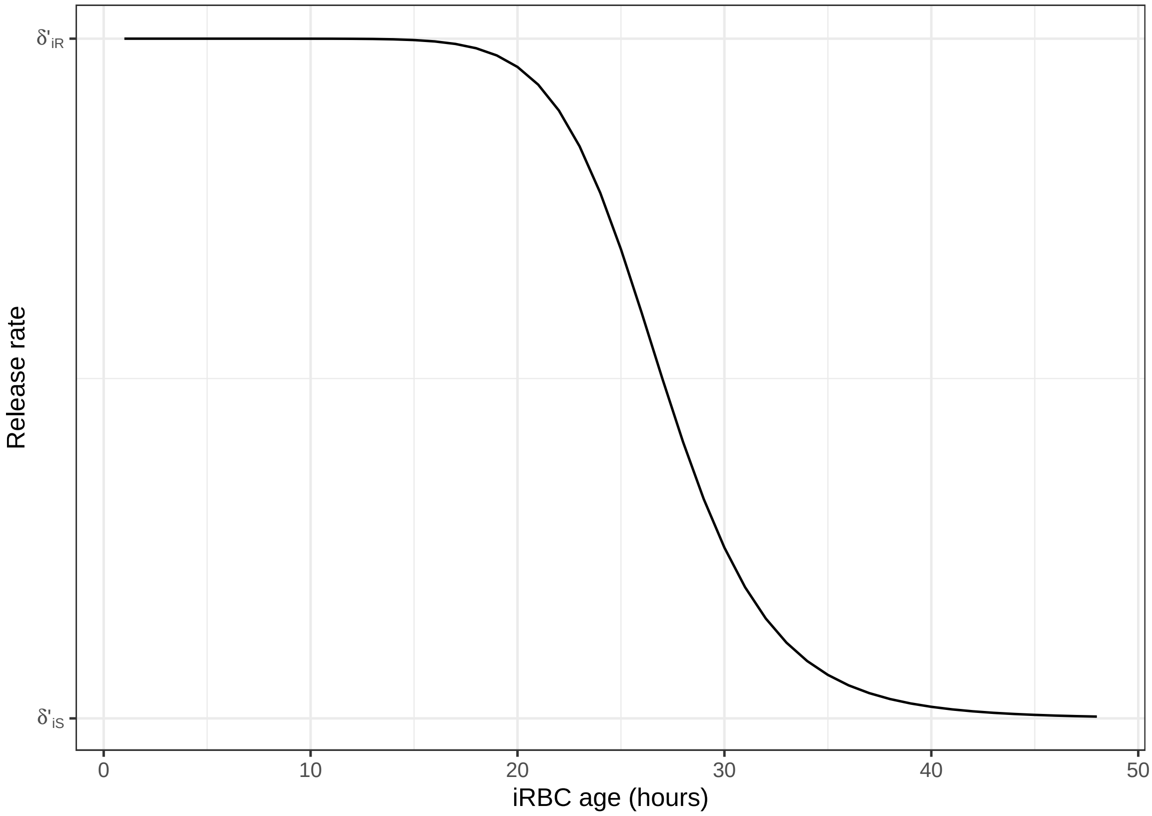


Figure S13: The iRBC release rate $\delta_i'(a)$ from the spleen into the circulation.

### S10 Infected RBC sequestration

The sequestration rate depends on the parameters $\zeta_{\text{sl}}$ and $\zeta_{50}$:

$$\zeta(a) = -\log(1 - 0.99) \cdot \frac{a^{\zeta_{\text{sl}}}}{a^{\zeta_{\text{sl}}} + \zeta_{50}^{\zeta_{\text{sl}}}} \tag{44}$$

Circulating iRBCs are sequestered at rate $\zeta(a)$, which is very low for Pf rings, and increases with maturity. The maximum rate corresponds to a sequestration probability of 0.99.

We denote the number of iRBCs sequestered in a time-step as $I_{c\to q}$:

$$I_{c\to q}(a,t) = I_c(a-1,t-1) \cdot (1 - \exp[-\zeta(a)]) \tag{45}$$

$$I_q(a,t) = I_q(a-1,t-1) + I_{c\to q}(a,t) \tag{46}$$

| Symbol | Description | Baseline value |
|---|---|---|
| $\zeta_{\text{sl}}$ | Slope parameter | 10 |
| $\zeta_{50}$ | Half-maximal age | 26 hours |

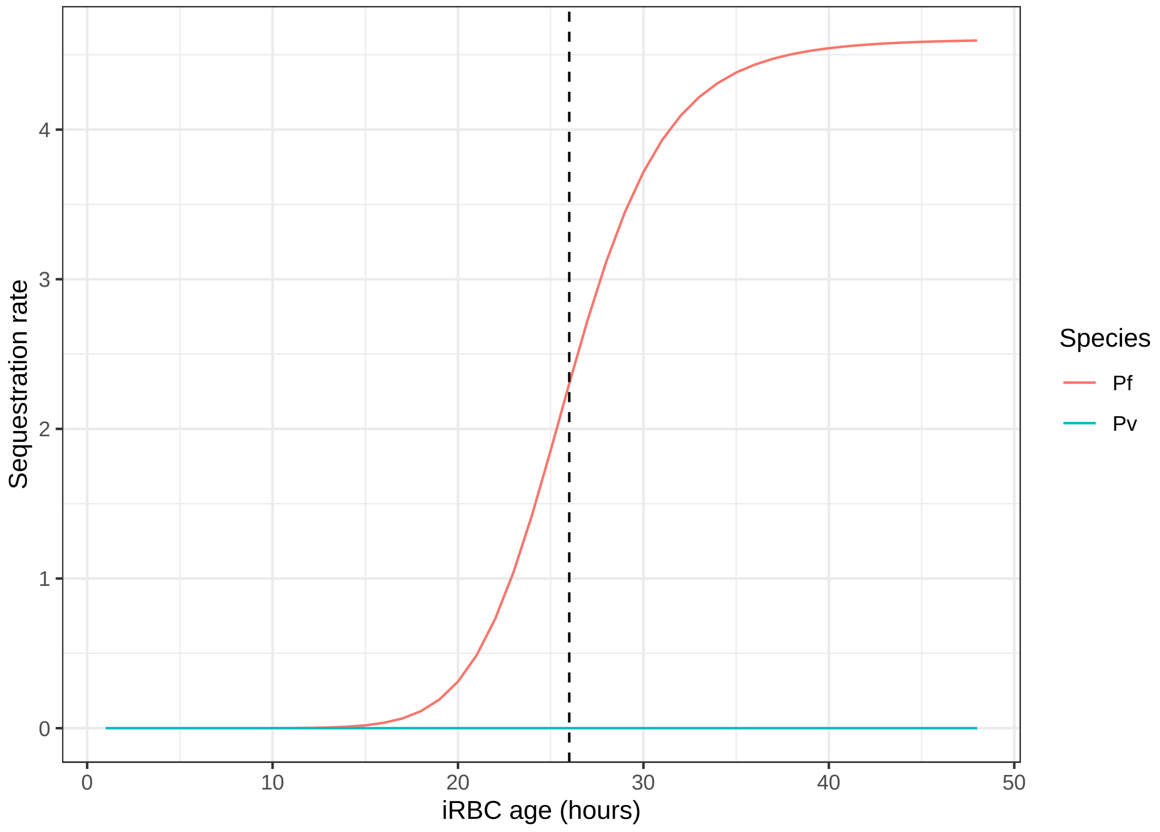


Figure S14: The age-specific rate $\zeta(a)$ of iRBC sequestration.

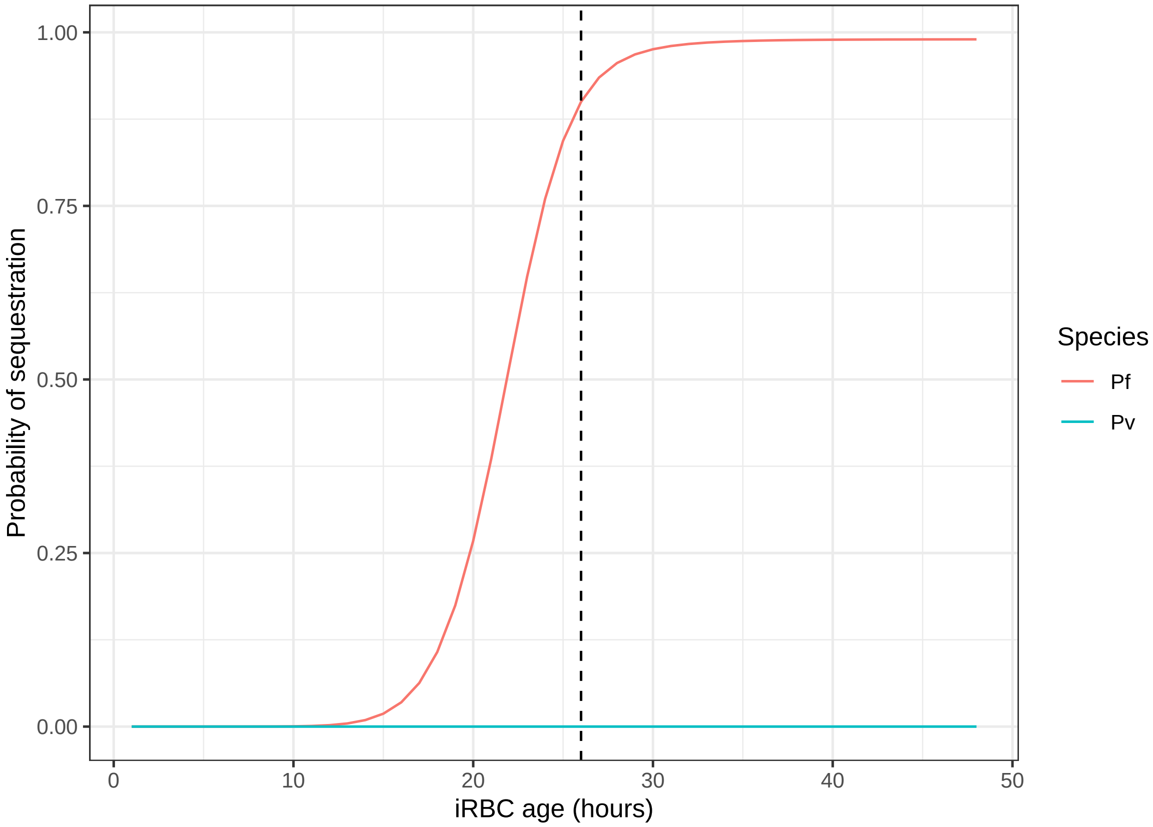


Figure S15: The age-specific probability of iRBC sequestration.

### S11 RBC destruction in the spleen

Phagocytosis rates depend on the macrophage population $M(t)$ and rate parameters $\lambda_u^{\text{sel}}$ and $\lambda_i^{\text{sel}}$:

$$\lambda_u(a,t) = \begin{cases} 0 & \text{for } a \leq T_M \\ \lambda_u^{\text{sel}} \cdot M_u(t-1) & \text{for } a > T_M \end{cases} \tag{47}$$

$$\lambda_i(a,t) = \lambda_i^{\text{sel}} \cdot M_i(t-1) \tag{48}$$

$$M_u(t) = M(t) \cdot \frac{\mathbf{U_s}(t-1)^{\gamma_M}}{\mathbf{U_s}(t-1)^{\gamma_M} + \mathbf{I_s}(t-1)^{\gamma_M}} \tag{49}$$

$$M_i(t) = M(t) \cdot \frac{\mathbf{I_s}(t-1)^{\gamma_M}}{\mathbf{U_s}(t-1)^{\gamma_M} + \mathbf{I_s}(t-1)^{\gamma_M}} \tag{50}$$

The macrophage population is defined with respect to the steady-state ratio of macrophages to RBCs retained in the spleen:

$$M(t) = k_M \left[ b_M \left( \mathbf{U_s}(t-1) + \mathbf{I_s}(t-1) \right) - M(t-1) \right] + M(t-1) \tag{51}$$

$$b_M = \frac{M_0}{U_{s,ss}} \tag{52}$$

Table S11: Model parameters for RBC destruction and macrophage populations.

| Symbol | Description | Baseline value |
|---|---|---|
| $\lambda_u^{\text{sel}}$ | Rate parameter for uRBCs | $5 \times 10^{-7}$ |
| $\lambda_i^{\text{sel}}$ | Rate parameter for iRBCs | $1.5 \times 10^{-11}$ |
| $k_M$ | Scaling factor | 0.01 |
| $b_M$ | Steady-state ratio of $M$ to $U_s$ | 0.0513979 |
| $\gamma_M$ | Scaling factor | 0.25 |
| $T_M$ | Age at which reticulocytes mature | 108 hours |

### S12 Differences between Pf and Pv

- The two species have different age-dependent merozoite preferences for uninfected RBCs; see RBC infection for details.
- Pf-infected RBCs can become sequestered in the microvasculature; see Infected RBC sequestration for details.

#### S13 Final RBC equations

We can now define the update rules for the RBC populations in the circulation and spleen, in terms of the equations defined above:

$$R_c(a,t) = \begin{cases} 0 & \text{for } a = 1 \\ R_{c\to c}(a,t) + U_{s\to c}(a,t) + r_{\to c}(a,t) - \nabla_{c\to c}(a,t) - \nabla_{s\to c}(a,t) & \text{for } 1 < a \leq T_M \end{cases} \tag{53}$$

$$N_c(a,t) = \begin{cases} R_{c\to c}(a,t) + U_{s\to c}(a,t) - \nabla_{c\to c}(a,t) - \nabla_{s\to c}(a,t) & \text{for } a = T_M + 1 \\ N_{c\to c}(a,t) + U_{s\to c}(a,t) - \nabla_{c\to c}(a,t) - \nabla_{s\to c}(a,t) & \text{for } T_M + 1 < a \leq T_U \end{cases} \tag{54}$$

$$U_c(a,t) = \begin{cases} R_c(a,t) & \text{for } 1 < a \leq T_R \\ N_c(a,t) & \text{for } T_R < a \leq T_U \end{cases} \tag{55}$$

$$U_s(a,t) = U_{s\to s}(a,t) + U_{c\to s}(a,t) - \nabla_{s\to s}(a,t) \tag{56}$$

$$I_c(a,t) = \begin{cases} \nabla_c(t) & \text{for } a = 1 \\ I_{c\to c}(a,t) + I_{s\to c}(a,t) & \text{for } 1 < a \leq T_I \end{cases} \tag{57}$$

$$I_s(a,t) = \begin{cases} \nabla_s(t) & \text{for } a = 1 \\ I_{s\to s}(a,t) + I_{c\to s}(a,t) & \text{for } 1 < a \leq T_I \end{cases} \tag{58}$$

where:

$$R_{c\to c}(a,t) = R_c(a-1,t-1) \cdot \exp[-\delta_u] \tag{59}$$

$$N_{c\to c}(a,t) = N_c(a-1,t-1) \cdot \exp[-\delta_u] \tag{60}$$

$$U_{s\to s}(a,t) = U_s(a-1,t-1) \cdot \exp[-\delta'_u - \lambda_u] \tag{61}$$

$$I_{c\to c}(a,t) = I_c(a-1,t-1) \cdot \exp[-\delta_i - \zeta] \tag{62}$$

$$I_{s\to s}(a,t) = I_c(a-1,t-1) \cdot \exp[-\delta'_i - \lambda_i] \tag{63}$$

### S14 Baseline outputs: no infection

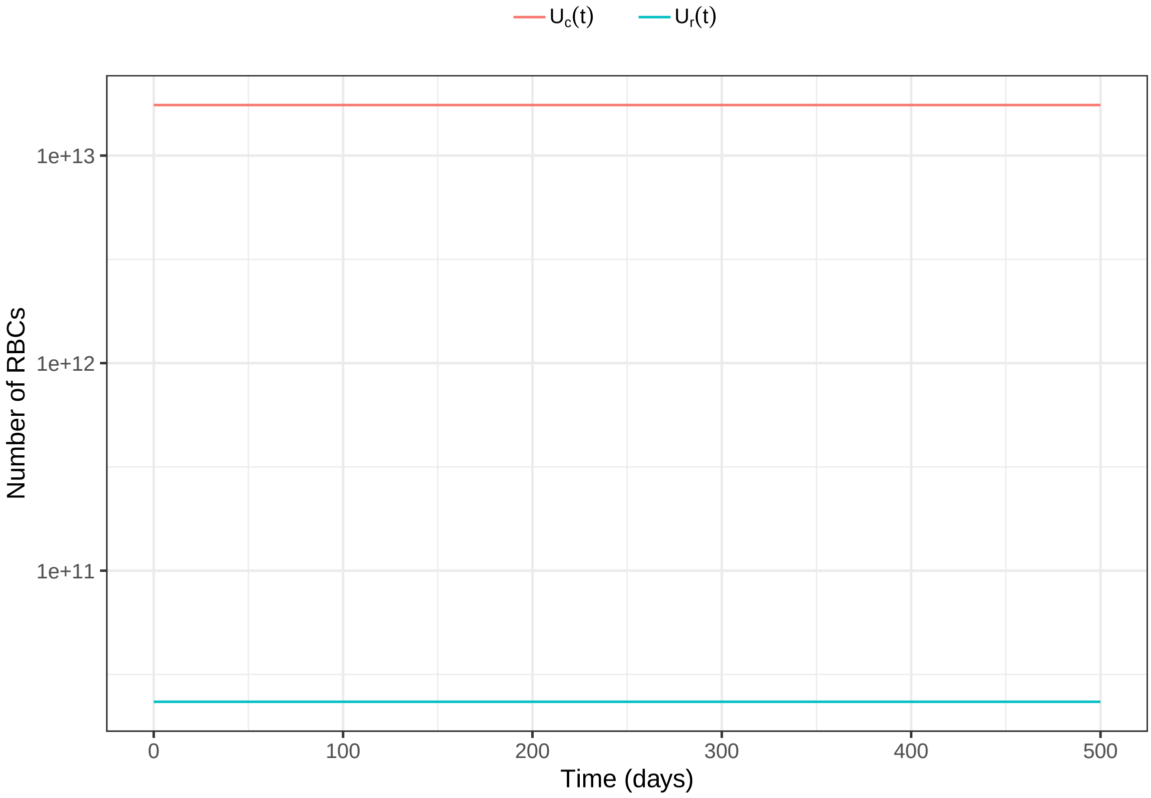


Figure S16: RBC populations over time for the baseline parameter values (no infection).

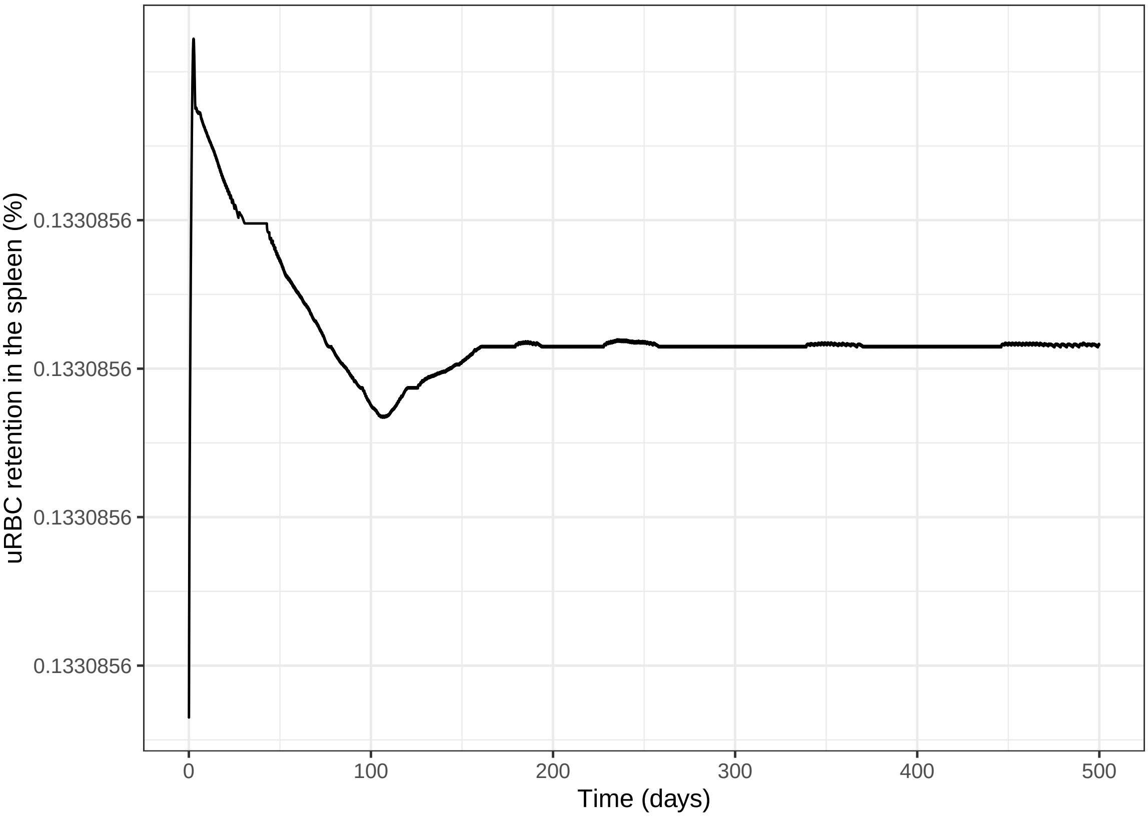


Figure S17: Uninfected RBC retention in the spleen (no infection).

#### S15 Baseline outputs: Pf

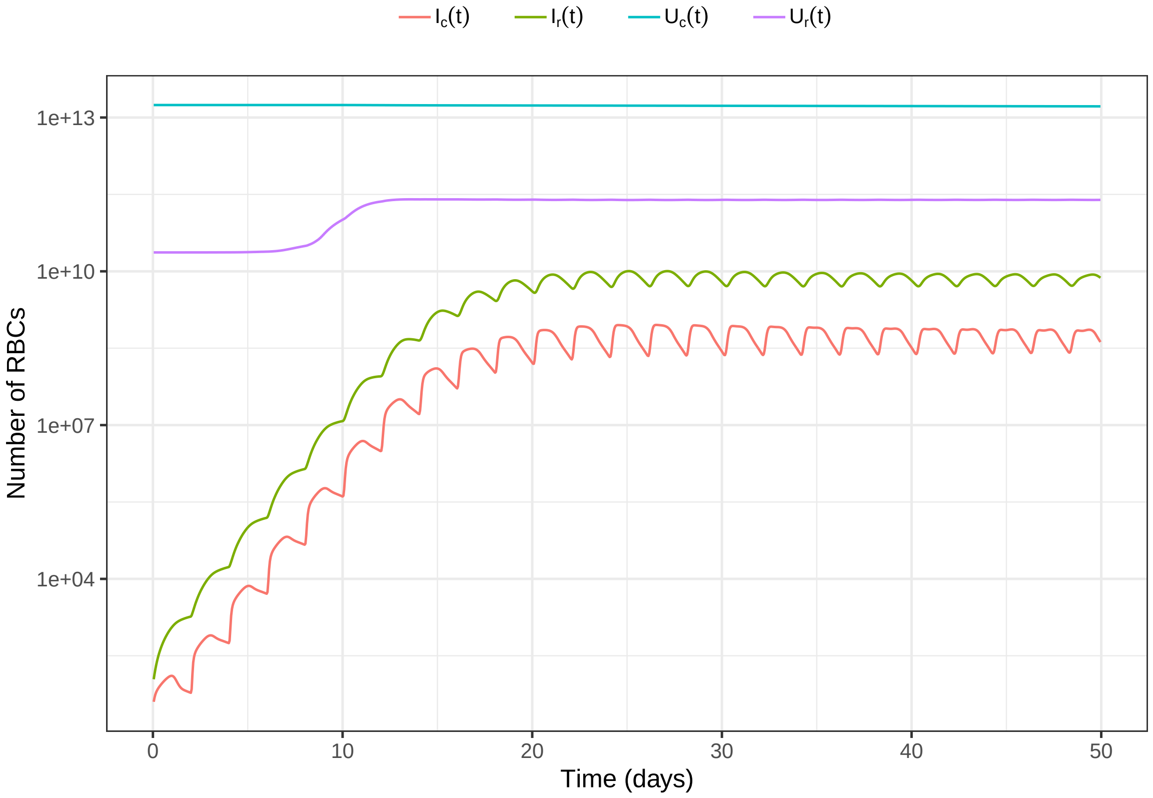


Figure S18: RBC populations over time for the baseline parameter values (Pf infection).

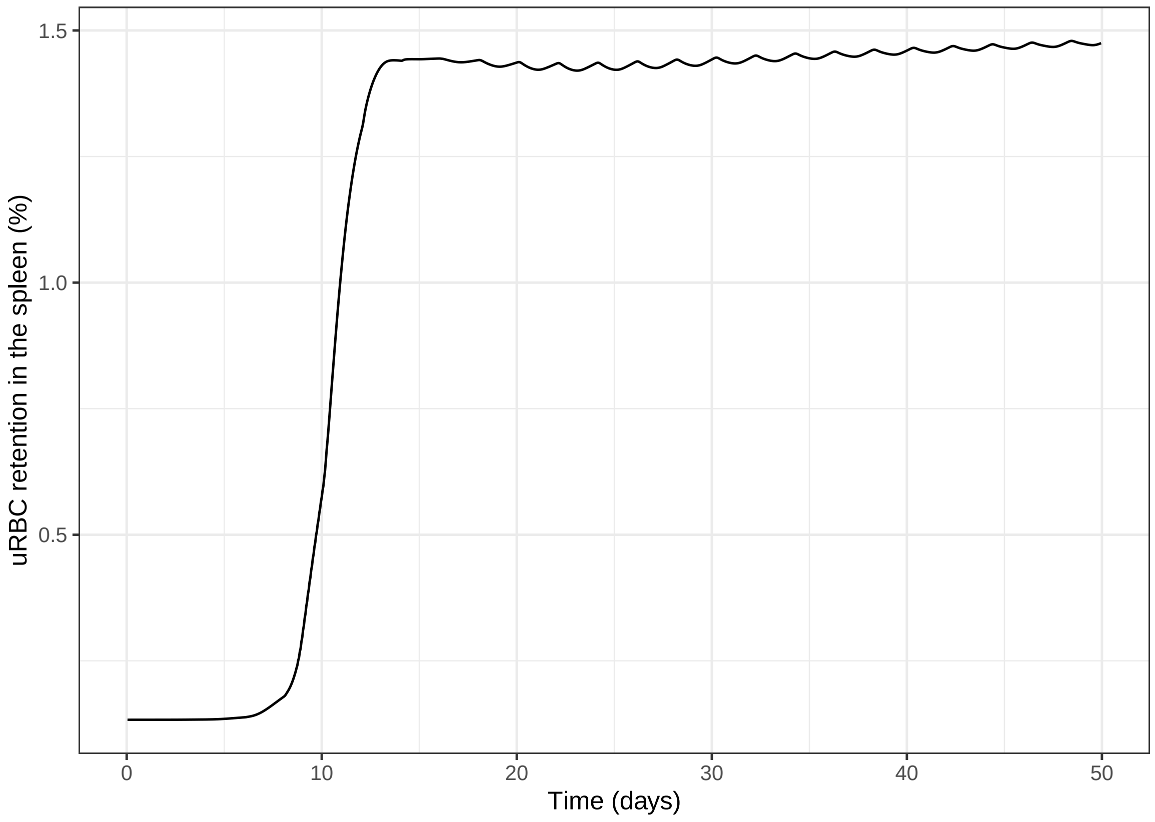


Figure S19: Uninfected RBC retention in the spleen (Pf infection).

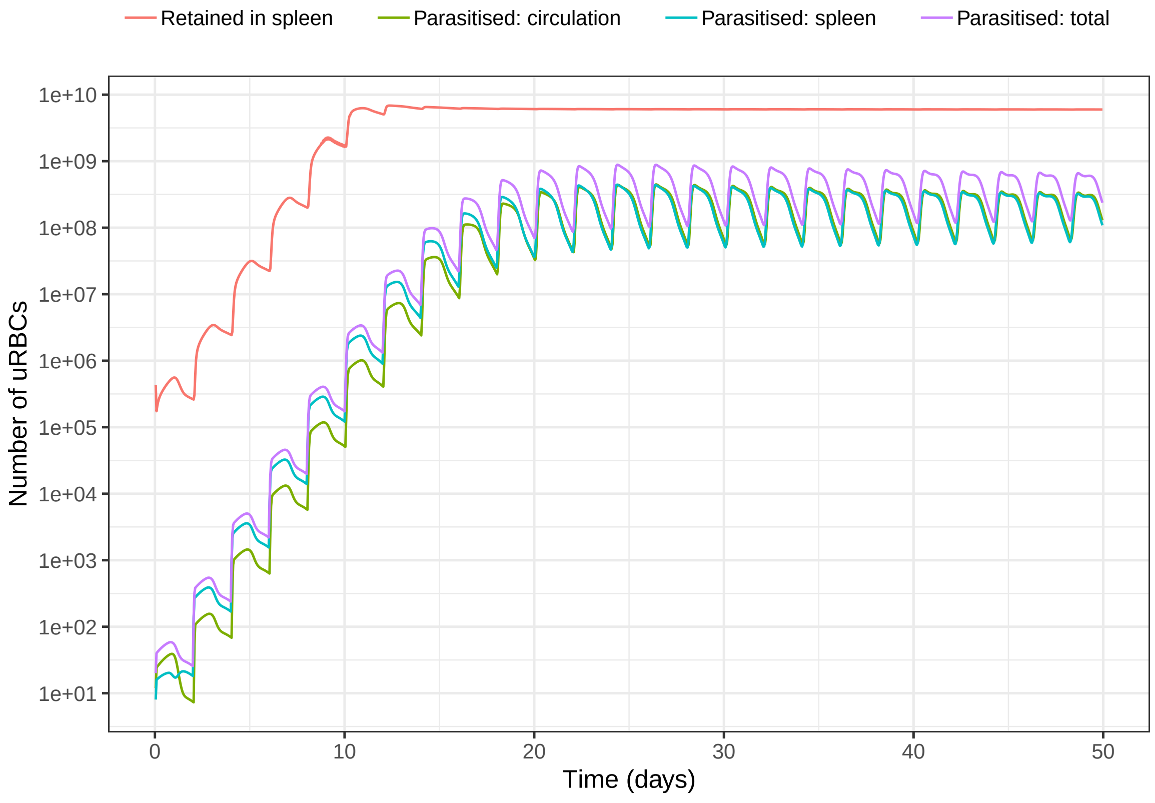


Figure S20: Uninfected RBC loss due to malaria, by infection and by retention in the spleen (Pf infection).

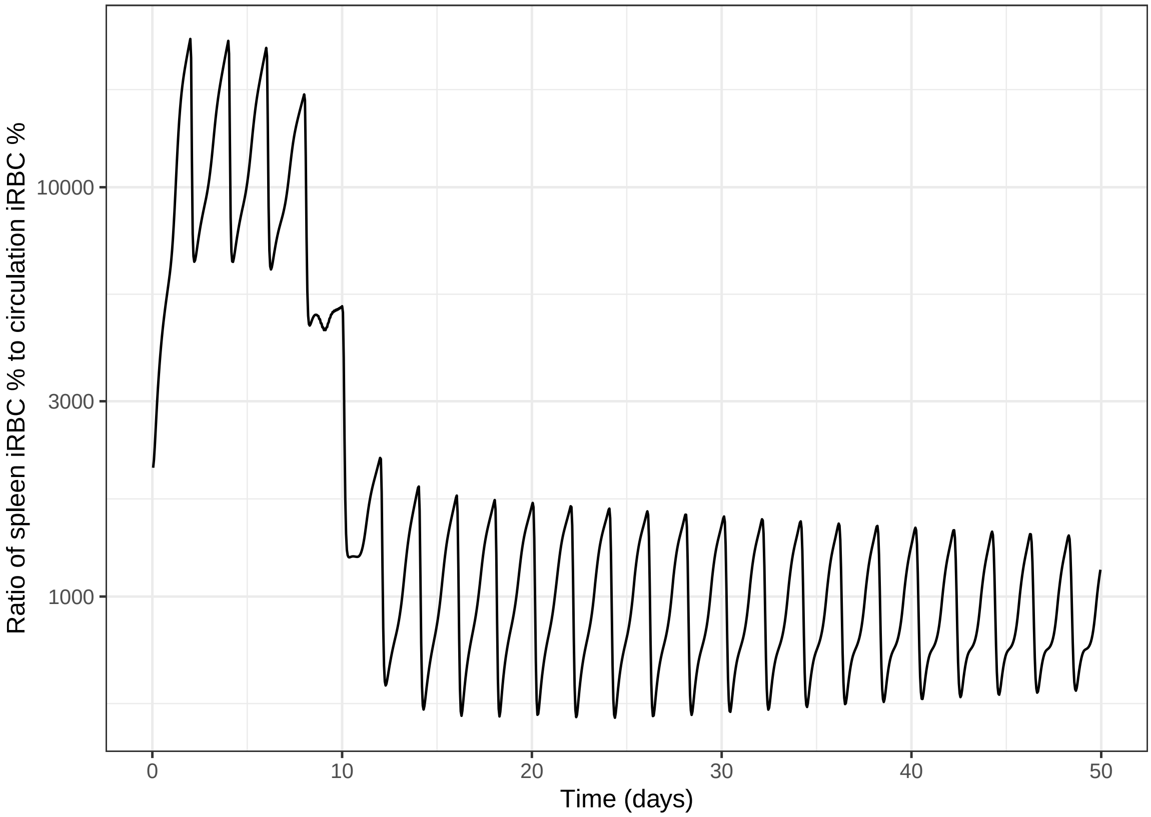


Figure S21: The ratio of (a) the proportion of RBCs in the spleen that are infected; to (b) the proportion of RBCs in the circulation that are infected (Pf infection).

### S16 Baseline outputs: Pv

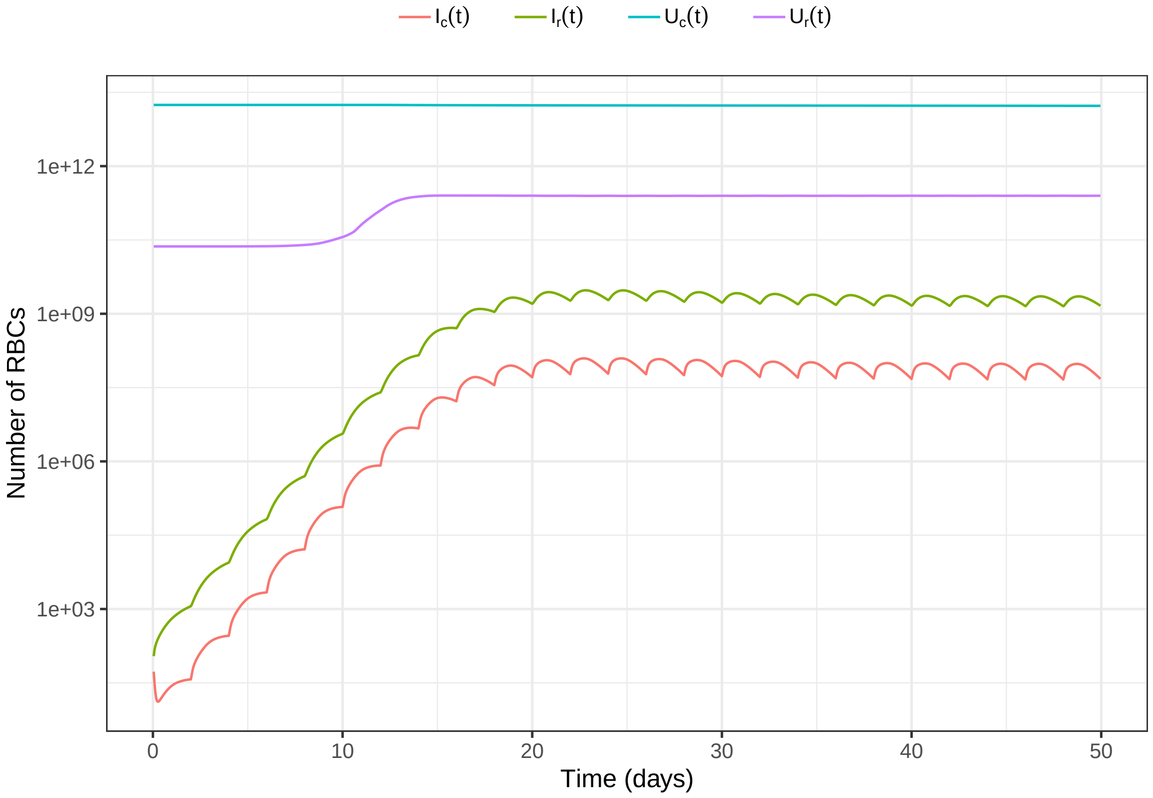


Figure S22: RBC populations over time for the baseline parameter values (Pv infection).

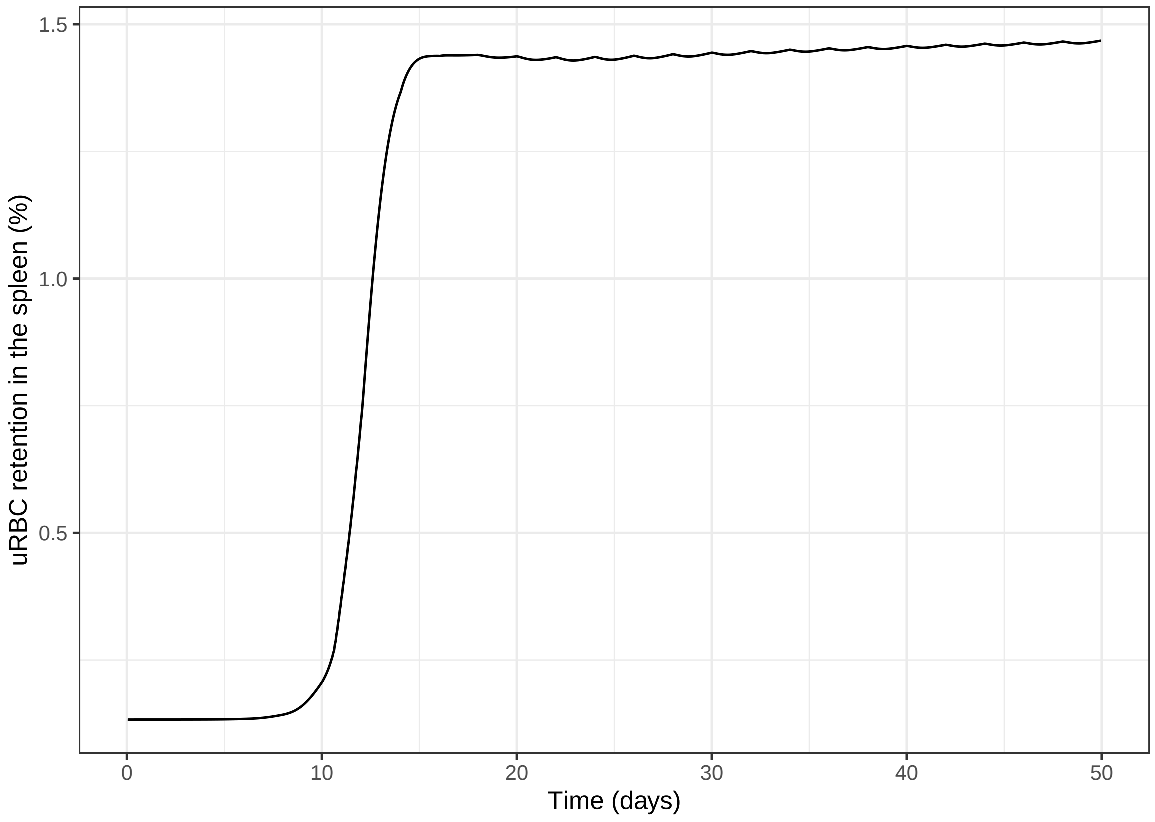


Figure S23: Uninfected RBC retention in the spleen (Pv infection).

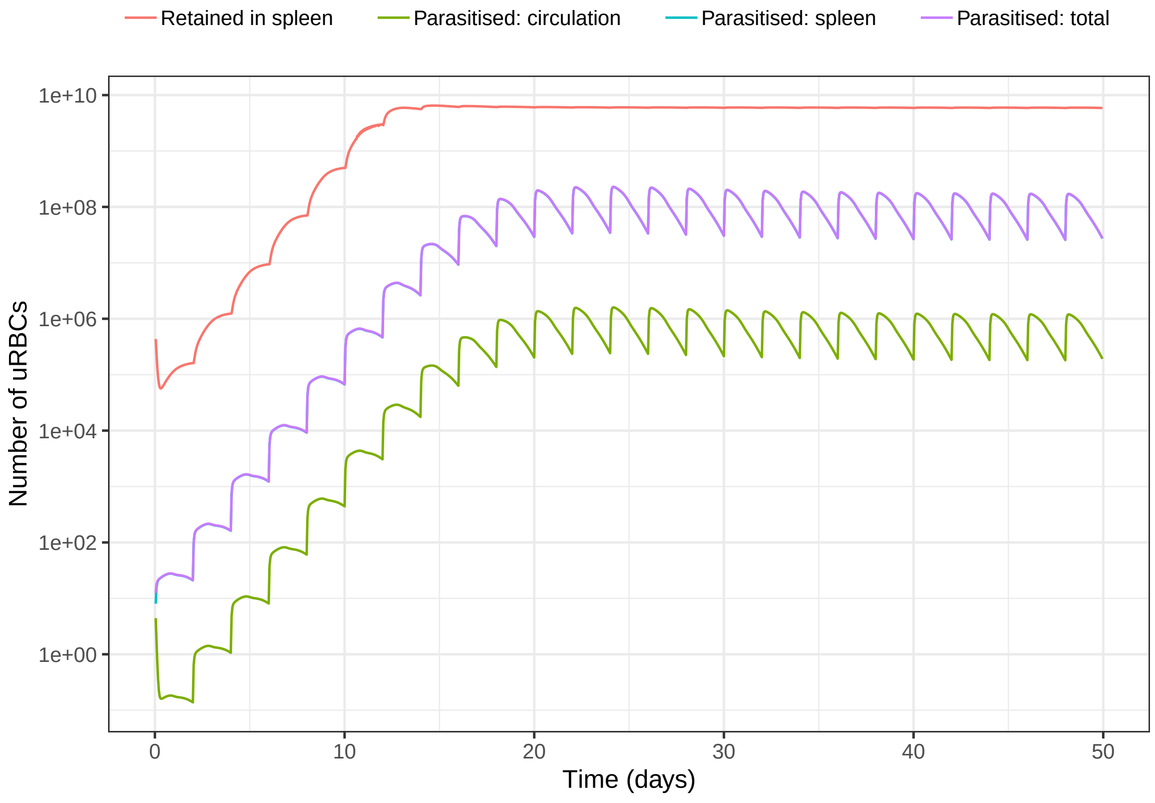


Figure S24: Uninfected RBC loss due to malaria, by infection and by retention in the spleen (Pv infection).

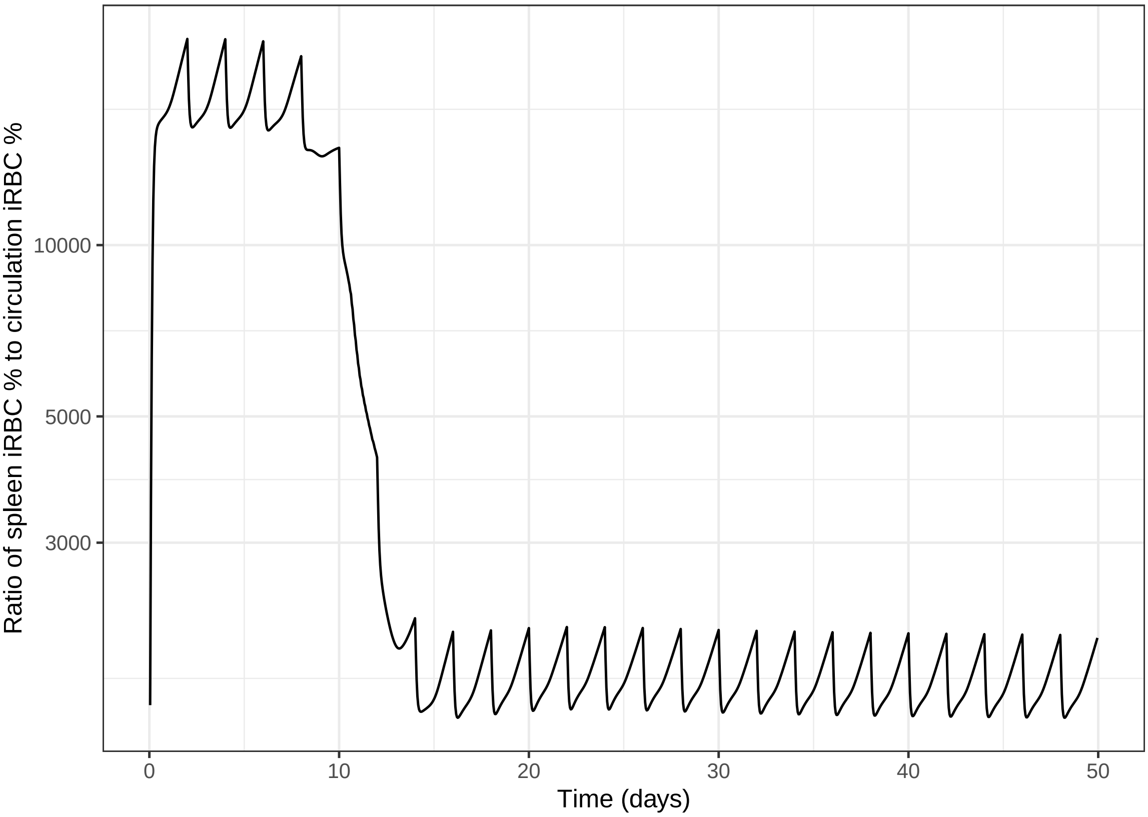


Figure S25: The ratio of (a) the proportion of RBCs in the spleen that are infected; to (b) the proportion of RBCs in the circulation that are infected (Pv infection).

#### S17 Model parameters

| Process | Parameter | Value |
|---|---|---|
| Normoblast production | $e_{\mathrm{sl}}$ | 16 |
| | $U_c^l$ | 0.33 |
| | $f_{\max}$ | 10 |
| Reticulocyte release | $\rho_0$ | 0.001 |
| | $\rho_s$ | 10 |
| | $\rho_i$ | 0.5 |
| | $T_R^{\min}$ | 24 |
| | $\kappa$ | $10^{-9}$ |
| uRBC removal | $\delta_U^A$ | 0.74303 |
| | $\delta_U^{\min}$ | $2.16405 \times 10^{-5}$ |
| | $\delta_U^{\max}$ | 1.206914 |
| | $\delta_U^{c50}$ | 2954.306 |
| | $\delta_U^g$ | 43.73335 |
| | $\nu$ | 0 |
| | $k_\nu^U$ | 1 |
| | $g_d^U$ | 1 |
| | $\delta_{50}^U$ | $10^{-7}$ |
| uRBC release | mag | 10 |
| | $\mu_U$ | 3.65 |
| | $\sigma_U$ | 0.0025 |
| RBC infection | $\omega$ | 0.1 |
| | PMF | 8 |
| iRBC removal | $\delta_i R$ (Pf) | 0.562 |
| | $\delta_i S$ (Pf) | 1.124 |
| | $\delta_i R$ (Pv) | 0.562 |
| | $\delta_i S$ (Pv) | 1.124 |
| | $\delta_I^{sl}$ | 10 |
| | $\delta_I^{c50}$ | 26 |
| | $k_\nu^I$ | 3 |
| | $\delta_{50}^I$ | $10^{-4}$ |
| iRBC release | $k_{iR}$ | 0.03 |
| | $k_{iS}$ | 0.01 |
| | $\delta'_{iR}$ | 0.01686 |
| | $\delta'_{iS}$ | 0.01124 |
| iRBC sequestration | $\zeta_{sl}$ | 10 |
| | $\zeta_{50}$ | 26 |
| RBC destruction | $\lambda_u^{sel}$ | $5 \times 10^{-7}$ |
| | $\lambda_i^{sel}$ | $1.5 \times 10^{-11}$ |
| | $k_M$ | 0.01 |
| | $b_M$ | 0.0513979 |
| | $\gamma_M$ | 0.25 |
| | $T_M$ | 108 |
